# Synergetic measures to contain highly transmissible variants of SARS-CoV-2

**DOI:** 10.1101/2021.11.24.21266824

**Authors:** Hang Su, Yafang Cheng, Christian Witt, Nan Ma, Ulrich Pöschl

## Abstract

**Background:** The public and scientific discourse on how to mitigate the COVID-19 pandemic is often focused on the impact of individual protective measures, in particular on vaccination. In view of changing virus variants and conditions, however, it seems not clear if vaccination or any other protective measure alone may suffice to contain the transmission of SARS-CoV-2.

**Methods:** Here, we investigate the effectiveness and synergies of vaccination and non-pharmaceutical interventions like masking, distancing & ventilation, testing & isolation, and contact reduction as a function of compliance in the population. Our new analysis accounts for the practical compliance in the population and for both droplet transmission and aerosol transmission.

**Findings:** For realistic conditions, we find that it would be difficult to contain highly contagious SARS-CoV-2 variants by any individual measure. Instead, we show how multiple synergetic measures have to be combined to reduce the effective reproduction number (*R*_e_) below unity for different basic reproduction numbers ranging from the SARS-CoV-2 ancestral strain up to measles-like values (*R*_0_ = 3 to 18). For *R*_0_ = 5 as reported for the Delta variant and ∼70% vaccination rate, the synergies of masking and distancing & ventilation with compliances around 30% appear sufficient to keep *R*_e_ < 1. In combination with 2-3 tests per week, this would work also at lower vaccination rates, e.g., in schools.

**Interpretation:** If the Omicron variant were to reach *R*_0_ = 8, it could still be contained with the synergetic measures outlined above. In case of measles-like transmissibilities (*R*_0_ = 12 to 18), higher compliances and testing rates or additional measures like general contact reductions would be required. The presented findings and approach can be used to design and communicate efficient strategies for mitigating the COVID-19 pandemic.

**Funding:** Max Planck Society.

**Research in context:** *Evidence before this study:* Studies on how to mitigate the COVID-19 pandemic are often focused on the impact of individual protective measures, in particular on vaccination. The effectiveness of non-pharmaceutical interventions (NPIs) like masking or distancing & ventilation are often under debate due to a lack of understanding of different transmission pathways (droplet versus aerosol transmission) and protective measures, in particular for the efficacy of masking and contrasting randomized trial results under different conditions (virus-limited vs. virus-rich) and at different levels of practical compliance. Thus, in view of more contagious variants such as Delta or Omicron, it is not clear if vaccination or any other protective measure alone may suffice to contain the transmission of SARS-CoV-2.

*Added value of this study:* Our analysis explicitly accounts for both droplet and aerosol transmission as well as for practical compliance in the population, which is the main reason for divergent results on the effectiveness of the same NPIs in different regions. This was not fully considered before and may have led to misunderstandings and misinformation about the actual effects of preventive measures. For realistic conditions, we find that it would be difficult to contain highly contagious SARS-CoV-2 variants by any individual measure. Instead, we show that combining multiple synergetic measures with realistic compliances can reduce *R*_e_ below unity without lockdown.

*Implications of all the available evidence:* Our findings and the presented scientific approach can be used to design and communicate efficient strategies for mitigating the COVID-19 pandemic for specific environments like schools as well as on a population level.

## Introduction

The COVID-19 pandemic has severe health, economic, and societal effects. Immunization by vaccination is one of the most important and prominent measures to control and mitigate the transmission of SARS-CoV-2, which has the benefit of not just reducing the transmission but also reducing the average severity of disease ^1^. Recent developments, however, suggest that the progress and effectiveness of vaccination may not suffice for suppressing or breaking waves of infection and swiftly mitigating the spread of COVID-19 ^2^. Besides vaccination, common further measures to control and contain the transmission of SARS-CoV-2 are universal masking, distancing & ventilation, contact reduction, and testing & isolation ^3-10^. Here, we investigate and quantify the effectiveness and synergies of these measures in reducing the effective reproduction number, *R*_e_. In the main text and figures we focus on a basic reproduction number of *R*_0_ = 5 that approximates the transmissibility of the Delta variant of SARS-CoV-2 ^11^. In the supplementary Figs, we also show results for *R*_0_ = 3 referring to the ancestral strain of SARS-CoV-2 and for *R*_0_ = 8 to 18 representing extremely contagious variants with transmissibilities similar to measles (Supplementary text S1). These high *R*_0_ scenarios may become relevant for designing measures and strategies against the newly emerging Omicron or further variants (Supplementary text S1).

A detailed account of the scientific approach and methods applied in our study is given in the Methods. Based on recent observations, we assume that the probability of SARS-CoV-2 transmission is on average reduced by approx. 70% for vaccinated persons ^12,13^ (Supplementary text S2). As illustrated in Fig. 1, universal masking reduces both the exhalation and the inhalation of respiratory viruses like SARS-CoV-2 (source control and wearer protection) and can thus reduce the probability of transmission by approx. 80% in case of surgical masks and approx. 99% in case of N95/FFP2 masks (Methods; Cheng et al., 2021). Physical distancing by at least 1-2 meters and proper ventilation of indoor environments can decrease the risk of transmission by respiratory droplets (> 0.1 mm) and aerosols (< 0.1 mm) by approx. 90% (Methods), whereby distancing primarily reduces droplet transmission and ventilation primarily reduces aerosol transmission ^3-6^. Reducing the number of contacts leads to a directly proportional decrease of *R*_e_, and the effects of testing & isolation of infected persons on *R*_e_ can be described as detailed in the Methods ^14^. Our investigations are focused on airborne transmission (including aerosols and droplets) rather than fomite transmission. Most of the investigated protective measures, however, would also reduce fomite transmission (vaccination, contact reductions, testing & isolation, masking via source control).

**Fig. 1.**
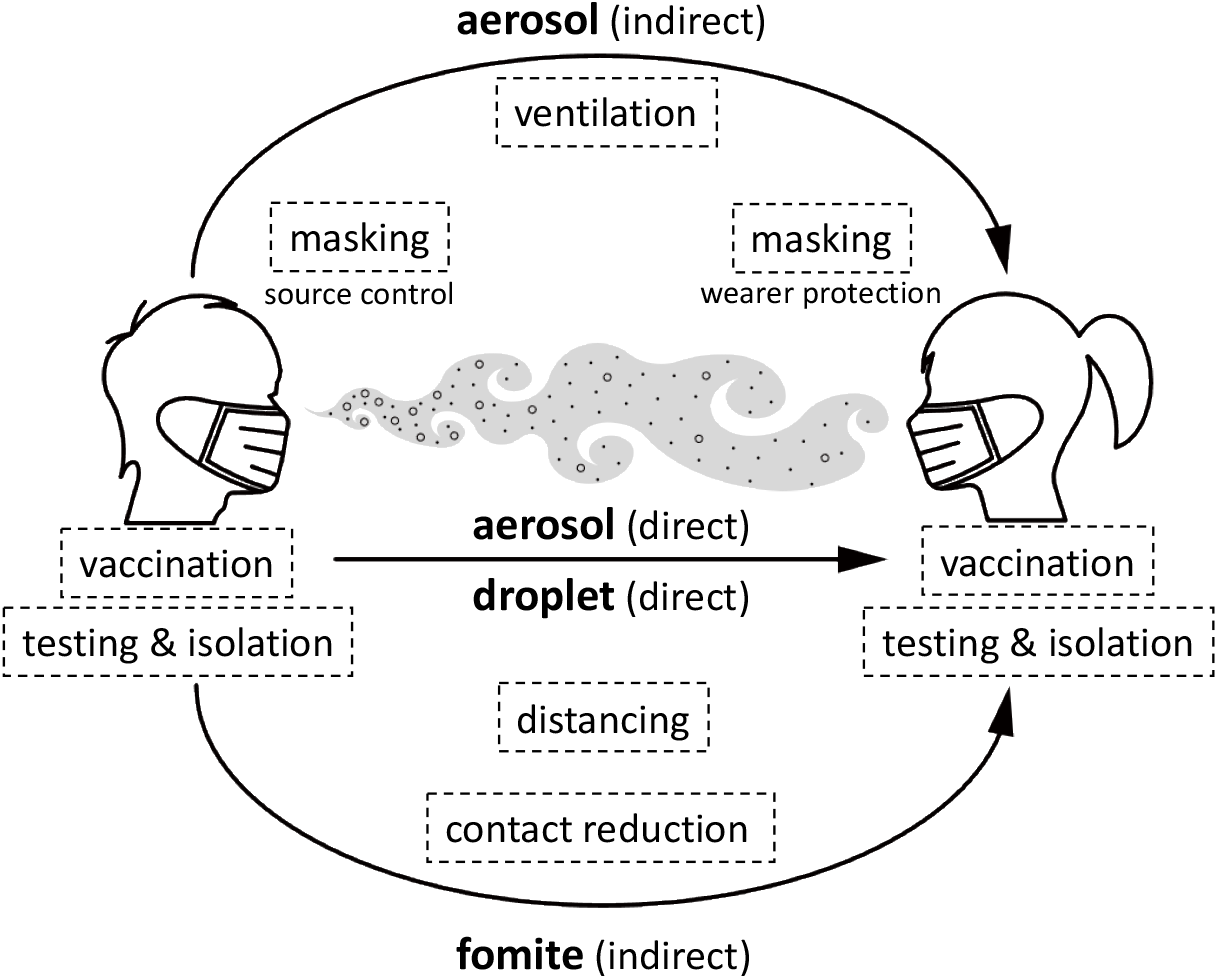
Schematic illustration of different transmission pathways and protective measures.

## Methods

In this study, we are building and extending the scientific approach developed in Cheng et al. (2021) ^5^. By definition, the basic reproduction number, *R*_0_, can be linked to the basic population average infection probability, *P*_0_, by

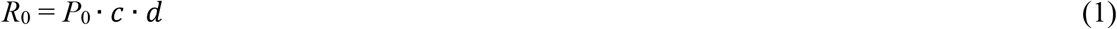

where *d* represents the average duration of infectiousness, and *c* represents the average daily number of human contacts (Supplementary text S1 and references therein). Vaccination, non-pharmaceutical interventions such as universal masking (surgical, N95/FFP2), distancing & ventilation, contact reduction, and testing & isolation can lead to reduced effective reproduction number, *R*_e_, by decreasing the infection probability *(P*), duration of infectiousness, or number of daily contacts.

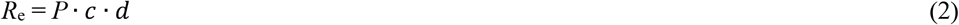

The effectiveness of an individual measure, *E*_i_, can be defined as

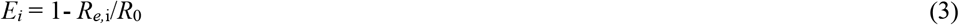

where *R*_*e*,i_ represents the effective reproduction number after implementing the measure *i*. The combined effectiveness of multiple independent measures, *E*_tot_, can be calculated by

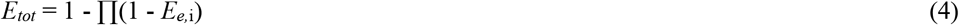

In the following, we described how the effects of different protective measures on *R*_e_ were calculated in this study.

The influence of vaccination on *R*_e_ is described *E*_vac_, which depends on the rate of vaccination, *V*_r_, and on the vaccine effectiveness against transmission, *E*_v_, i.e., the average degree by which the probability of SARS-CoV-2 transmission is reduced in the vaccinated population:

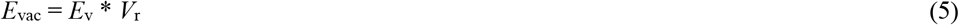

Universal masking is an effective measure against airborne transmission of SARS-CoV-2 ^5,10,15,16^ (Supplementary text S3). The influence of masking on *R*_e_ is described by *E*_mask_, which is calculated for both aerosol transmission (via respiratory particles with diameters < 0.1 mm) and droplet transmission (via respiratory droplets with diameters > 0.1 mm). The effects of masking on aerosol transmission of SARS-CoV-2 depends on *R*_0_ (reflecting population average infection probability) and have been calculated the same way as in Cheng et al. (2021) ^5^. Because the effectiveness of masking depends on the variability of infection probability (Eq. 1 & 2), we adopted a standard deviation of *σ* = 2 to describe the virus load distribution ^5^. The effect of compliance with masking, *m*, on the infection probability via aerosol transmission, *P*_mask,a_, can be described by Eq. 6 ^5^:

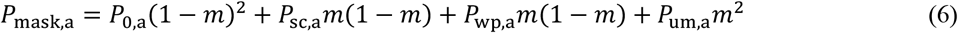

Here *P*_0,a_ represents the population average infection probability via aerosol transmission when no one wears masks; *P*_sc,a_ represents the average infection probability in the case of source control (mask wearing by infected persons); *P*_wp,a_ represented the average infection probability in the case of wearer protection (mask wearing by susceptible persons); and *P*_um,a_ represent the average infection probability in the case of universal masking as determined by Cheng et al. (2021) ^5^. Similarly, we can calculate the effects of universal masking on droplet transmission by

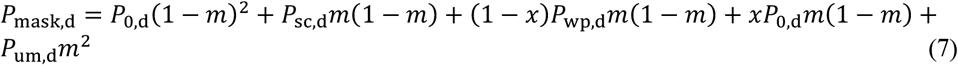

With regard to wearer protection against droplet transmission, the additional terms in Eq. 7 take into account that droplet transmission may also occur through the eyes of the mask wearer ^17^. The fractional contribution of droplet transmission through the eyes (eye infection) is described by the parameter *x*, and the fractional contribution of droplet transmission through mouth and nose is given by (1-*x*). By inserting *P*_mask,a_ or *P*_mask,d_ in Eq. 2 and 3, we calculated a mask effectiveness for aerosol transmission, *E*_mask,a_, and a mask effectiveness for droplet transmission, *E*_mask,d_. Because the relative contributions of aerosol and droplet transmission have not yet been determined, we took the smaller value for *E*_mask_ to obtain a conservative estimate for the overall effectiveness of masking. For *x* = 0.3 and as illustrated in Fig. S2, this implies *E*_mask_ = *E*_mask,a_ for surgical masks and *E*_mask_ ≈ *E*_mask,a_ ≈ *E*_mask,d_ for N95/FFP2 masks.

For proper physical distancing by at least 1-2 m, we assume an effectiveness of approximately ∼90% against direct transmission by droplets and aerosols, corresponding to the respiratory particle volume fraction typically lost through gravitational settling over such distances (Supplemental text S4, Fig. S3) ^7,19^. In contrast to physical distancing, regular ventilation has little effect on direct transmission by semi-ballistic respiratory droplets (> 0.1 mm) but reduces indirect transmission by equilibrated respiratory aerosols (< 0.1 mm) in indoor environments both in the near-field and the far-field ^5,6,18^. Changing from passive ventilation to high rates of active ventilation can reduce the airborne virus concentration and probability of indirect transmission by aerosols with an effectiveness up to ∼90% ^5^ (Supplemental text S4). Thus, we assume an effectiveness of *E*_d&v_ ≈ 90% for the combined influence of distancing & ventilation on *R*_e_.

The influence of testing & isolation on *R*_e_ can be described by an effectiveness of

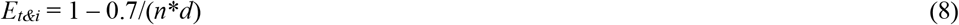

where *n* is the number of tests per week and *d* is the duration of infectiousness. The basis for this equation and how to consider relevant parameters like the average duration of infectiousness or false negative rates of testing are detailed in the supplement (Supplementary text S5).

## Results

### Individual measures and synergetic effects

For each of the investigated protective measures, Figure 2 shows how *R*_e_ decreases with increasing compliance in the population. Vaccination alone (black line) can reduce the reproduction number from *R*_0_ = 5 to *R*_e_ = 2.5 at 70% compliance, which corresponds approximately to the current rate of vaccination in Germany (https://impfdashboard.de/). Even at 100% compliance, however, vaccination alone would not reduce *R*_e_ below 1 as required to contain the transmission. Without other protective measures, *R*_e_ would remain as high as 1.5, leading to continued exponential growth. In other words, the currently available vaccines are highly protective against the disease and severe outcomes of COVID-19 ^12,13^, but they are not sufficient to contain and end the transmission of SARS-CoV-2 without synergetic measures. For *R*_0_ = 5 or higher basic reproduction rates, even a vaccine that reduces the probability of infection and transmission by 95% ^1^ would require vaccination rates higher than 85% to decrease *R*_e_ below 1 (Fig. S1). In theory, distancing & ventilation alone (yellow line) could decrease *R*_e_ to 0.5 at 100% compliance, but at more realistic compliance rates around 50% as discussed below, *R*_e_ would also remain above 1. Similarly, universal masking (red line) with 100% compliance could bring *R*_e_ close to 1 in case of surgical masks (Fig. 2A) and well below 1 in case of N95/FFP2 masks (Fig. 2B), but at more realistic compliance rates around 50%, *R*_e_ would again remain above 1.

**Fig. 2.**
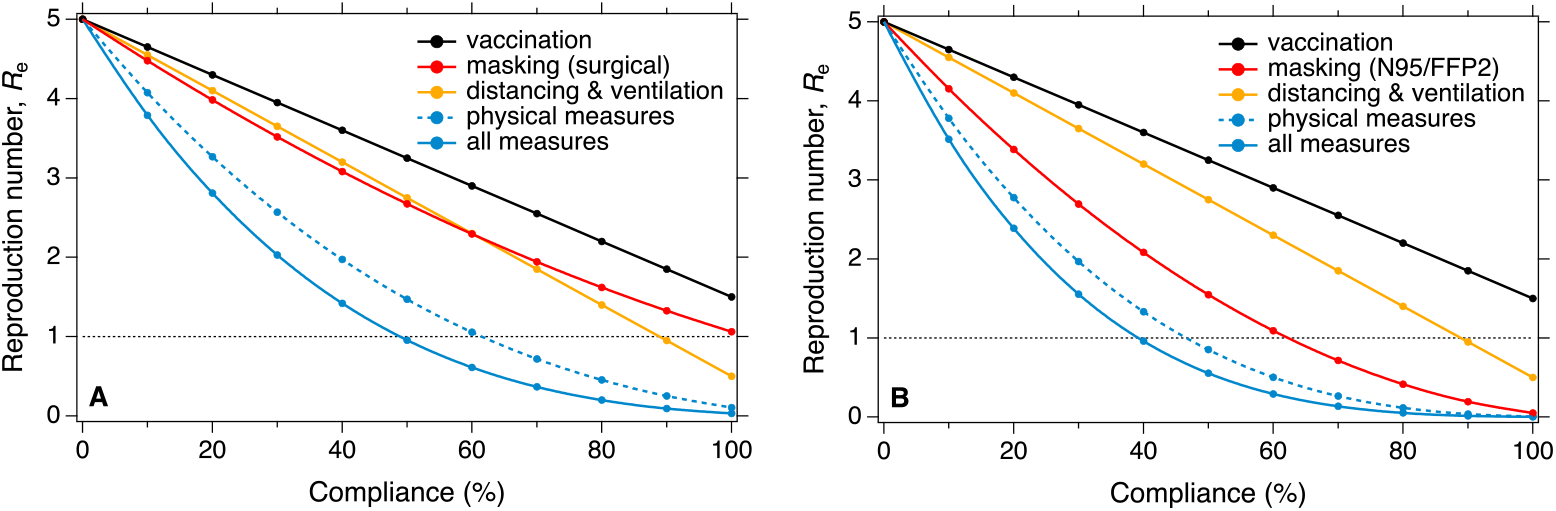
Effectiveness of individual and combined measures. Reduction of effective reproduction number, *R*_e_, as a function of compliance with different protective measures for a basic reproduction number *R*_0_ = 5 approximately reflecting the transmissibility of the Delta variant of SARS-CoV-2. Panels **A** and **B** refer to universal masking with surgical masks and N95/FFP2 masks, respectively. The curve labeled “physical measures” refers to the combination and synergy of universal masking plus distancing and ventilation; the curve labeled “all measures” refers to the combination and synergy of the physical measures with vaccination.

When all these measures are combined (solid blue line), compliance rates around 50% are sufficient to bring *R*_e_ close to 1 in case of surgical masks (Fig. 2A) and well below 1 in case of N95/FFP2 masks (Fig. 2B). The steep non-linear decay of the “all measures” curve and the very low *R*_e_ values obtained at high compliance highlight the strong synergetic effect that results from combining multiple protective measures and multiplying their individual effects. Even if only universal masking were combined with distancing & ventilation (“physical measures”, dashed blue line), *R*_e_ would fall below 1 at 50% compliance with N95/FFP2 masking (Fig. 2B). Note, however, that 50% compliance with masking are not easy to achieve as discussed below and in Cheng et al. (2021). We are not suggesting to promote these physical measures without vaccination, which would also be missing the benefit of reducing both the transmission of the virus and the severity of the disease by immunization ^1,12,13^. Nevertheless, the “physical measures” curve shows, that the synergetic effects of combining and properly applying these simple measures are strong enough to reduce the reproduction number substantially, e.g., for breaking or suppressing waves of infection.

The actual rates of vaccination vary widely due to different supplies, age limits, and willingness. For example, the percentages of fully vaccinated people are around 39.4% in India, 61.4% in the U.S.A., 70.2% in Germany, 87.8% in Portugal, and 46.6% worldwide at this time (21 December 2021) ^23^. In our study, we are not explicitly accounting for persons immunized by recovery from the disease. Depending on the level of immunization, they can be implicitly included in the vaccination rate (compliance). Given an approximate efficacy of 70% and an approximate upper limit of 90% for compliance, vaccination can only reduce *R*_e_ from 5 to approx. 1.9. Recent observations suggest that the efficacy of currently available vaccines against the transmission of the new Omicron variant is lower and may thus contribute less to reducing the effective reproduction number of SARS-CoV-2 (Supplemental text S2).

For universal masking, 100% compliance would be difficult to achieve because masking is not always possible and practical, for example at home, during eating or drinking in restaurants and bars, in schools and kindergartens, etc. ^5^. The potential importance of such situations is demonstrated by a recent modeling study attributing around 10% to 40% of daily infections to restaurants and cafés/bars ^24^. Moreover, a lack of willingness to follow recommendations or mandates for mask use may also lead to low compliance with mask wearing. For example, inpatient respiratory protection studies show that adherence rates vary from 10% to 84% for health care personnel ^25^. Similar effects can be expected when wearing masks with low efficiency or poor fit and high penetration or leakage rates ^5,26^. Combining these effects, we may estimate ∼50% as an effective upper bound for the compliance with universal masking (Supplementary text S3). For physical distancing, we may expect a similar effective upper limit of compliance because distancing may be difficult under the same or similar conditions that are unfavorable for masking. With regard to ventilation, earlier investigations indicate that the ventilation of indoor environments is often much lower than recommended, and the values recommended for common indoor environments are also lower than the ventilation rates used for effective infection control in health care units ^6,27^. We found no data specifically suited for estimating population-average ventilation effects ^6^, but based on the available literature we assume that the effective upper limit of compliance with distancing & ventilation is similar to the value estimated for distancing (approx. 50%).

Thus, it would be difficult to contain SARS-CoV-2 transmission and end the pandemic by any individual measure as currently available under realistic conditions. On the other hand, Fig. 2 shows that the synergetic effects of combining the investigated protective measures at realistic levels of compliance can decrease the effective reproduction number from *R*_0_ = 5 as reported for the Delta variant of SARS-CoV-2 to well below 1. In case of new virus variants such as Omicron, the efficacy of vaccines may be reduced, but the effectiveness of simple physical measures should not change much. In case of higher basic reproduction numbers up to measles-like transmissibilities, higher compliances or additional measures would be required as illustrated in Fig. S4 for *R*_0_ = 8, 12 and 18, respectively, and further discussed below. In the following, we explore the synergies of combining vaccination with universal masking, distancing & ventilation, contact reduction, and testing & isolation.

### Combining vaccination with non-pharmaceutical interventions

Figures 3A and 3B show how *R*_e_ decreases as a function of compliance with universal masking for different vaccination rates in the population, assuming that vaccine efficacy against virus transmission is 70% and that no other protective measures are applied. The basic reproduction number *R*_0_ is 5, and the reduced starting values of *R*_e_ at zero compliance with masking correspond to different vaccination rates. At a vaccination rate around 70% (black line), decreasing *R*_e_ below 1 would require compliances higher than 70% for surgical masking (Fig. 3A) and higher than 50% for N95/FFP2 masking (Fig. 3B). In practice, such compliance values appear unrealistically high as discussed above, indicating that controlling the transmission of SARS-CoV-2 in a population with moderate vaccination rate (e.g., in Germany) requires additional measures beyond vaccination and masking. At a vaccination rate around 90% (yellow line), decreasing *R*_e_ below 1 would require compliances around 50% for surgical masking (Fig. 3A) and 30% for N95/FFP2 masking (Fig. 3B). Thus, high compliance with universal masking may suffice to prevent or suppress potential waves of infection in populations with high vaccination rates (e.g., in Portugal).

**Fig. 3.**
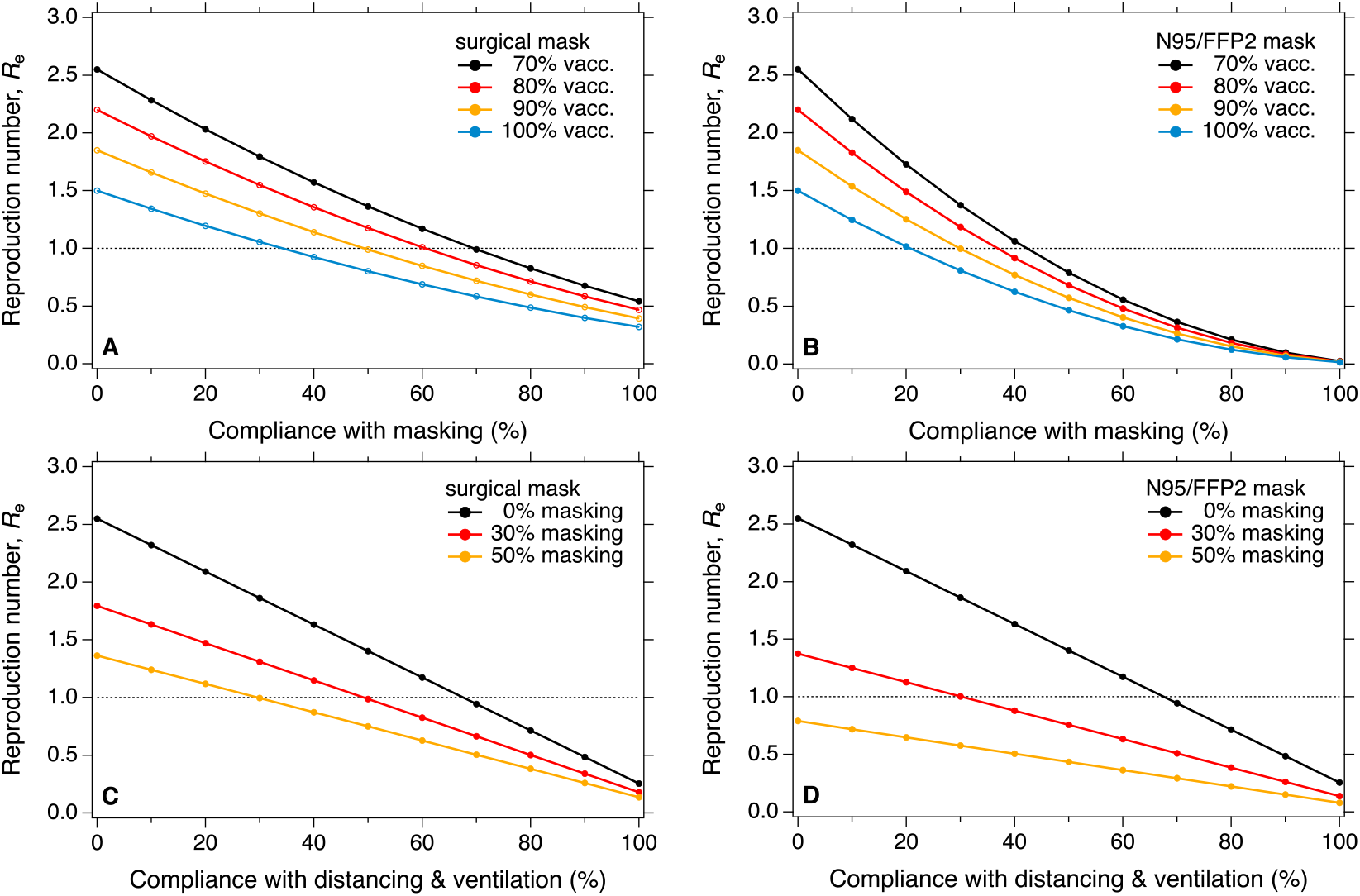
Effectiveness of masking and distancing & ventilation for different vaccination rates and compliances. Reduction of effective reproduction number, *R*_e_, as a function of compliance with universal masking for different vaccination rates (**A, B**) and as a function of compliance with distancing & ventilation for different levels of masking at a fixed vaccination rate of 70% (**C, D**). Basic reproduction number *R*_0_ = 5; reduced starting values of *R*_e_ at zero compliance with masking correspond to different vaccination rates. Left column corresponds to universal masking with surgical masks, right column corresponds to N95/FFP2 masks.

In Figs. 3C and 3D, we include the effect of distancing & ventilation. They show how *R*_e_ decreases as a function of compliance with distancing & ventilation for different compliances with universal masking at a fixed vaccination rate of 70%. Different starting values of *R*_e_ at zero compliance reflect the effects of vaccination and different levels of masking. Without masking (black line), decreasing *R*_e_ below 1 would require compliances higher than 70%, which appear unrealistically high as discussed above. With universal masking at a level of 30% (red line), decreasing *R*_e_ below 1 would require distancing & ventilation compliances around 50% in case of surgical masks (Fig. 3C) and around 30% for N95/FFP2 masks (Fig. 3D). As discussed above, compliance levels around 30% are not unrealistic. Thus, high compliance with universal masking in combination with distancing & ventilation may suffice to prevent or suppress waves of infection in populations with moderate vaccination rates (e.g., in Germany).

In Fig. S5, we include the effect of contact reduction. It shows how *R*_e_ decreases as a function of contact reduction for different compliances with universal masking at a fixed vaccination rate of 70%. With universal masking at 30% compliance (Figs. S5A and S5B, red line), decreasing *R*_e_ below 1 would require contact reductions by approx. 50% in case of surgical masks and approx. 30% for N95/FFP2 masks. According to recent studies ^8,28-30^, 30% contact reduction would correspond to partial confinement and 50% would correspond to a lockdown. When universal masking is combined with distancing & ventilation at 30% compliance (Figs. S5C and S5D, red line), contact reductions by approx. 30% would decrease *R*_e_ slightly below 1 for surgical masks and as low as 0.7 for N95/FFP2 masks. Thus, moderate contact reductions (around 30%, partial confinement) combined with distancing & ventilation and N95/FFP2 masking at moderate levels of compliance (around 30%) may suffice to prevent infection waves. The synergetic effects of contact reduction combined with other protective measures are further illustrated in Figs. S6, S7 and S8, including higher basic reproduction numbers (*R*_0_ = 8, 12 and 18, respectively). In case of measles-like transmissibilities (*R*_0_ = 12 to 18), even high levels of compliance (∼50%) with masking and distancing & ventilation plus vaccination (∼70%) would not suffice, and additional measures like general contact reductions (∼50%) would be required to bring *R*_e_ below 1 (Figs. S11 and S12).

Testing & isolation of infected persons is a protective measure particularly common and relevant for mitigating the transmission of SARS-CoV-2 in schools ^14^. Figure 4 shows a pronounced non-linear dependence of *R*_e_ on the frequency of testing (number of tests per week) for different vaccination rates and compliances with universal masking and distancing & ventilation. The latter are key measures and essential tools to contain the transmission in schools and keep them operational during the pandemic. Even for largely unvaccinated groups such as primary school children, *R*_e_ can be kept as low as 0.5-0.9 by 2-3 tests per week combined with distancing & ventilation and surgical or N95/FFP2 masking at moderate compliance levels around 30% (red lines, Figs. 4A and 4B). At a vaccination rate of 70%, similar results can be achieved just by testing and masking (red lines, Figs. 4C and 4D). At 70% vaccination rate, 30% masking, and 30% distancing & ventilation, even one test per week may suffice to keep *R*_e_ below 1 and contain the spread of SARS-CoV-2 (red lines, Figs. 4E and 4F). Figure S9 shows the results obtained for various further combinations of protective measures at different levels of compliance. In practice, the frequency of testing has to be adjusted according to the rates of false negative results ^22^ (Methods), and the effects of incomplete isolation have to be considered, which may reduce the effectiveness of this measure. Nevertheless, testing & isolation may be highly effective not only in educational but also in workplace and private environments, especially with increasing vaccination rates (see Figs. S9E and S9F). Testing & isolation combined with all other available measures and high compliances may become essential for containing SARS-CoV-2 transmission in educational and workplace environments if more contagious variants emerge (see Figs. S10, S11 and S12 for *R*_0_ = 8, 12 and 18, respectively). In case of measles-like transmissibilities (*R*_0_ = 12 to 18) and low efficacy of vaccination against transmission, high practical compliance with all NPIs including general contact reductions (∼50%) plus 3-4 tests per week could still bring *R*_e_ well below unity (< 0.7) and contain, if not terminate, the spread of SARS-CoV-2 in a population (Figs. S11E and S12E).

**Fig. 4.**
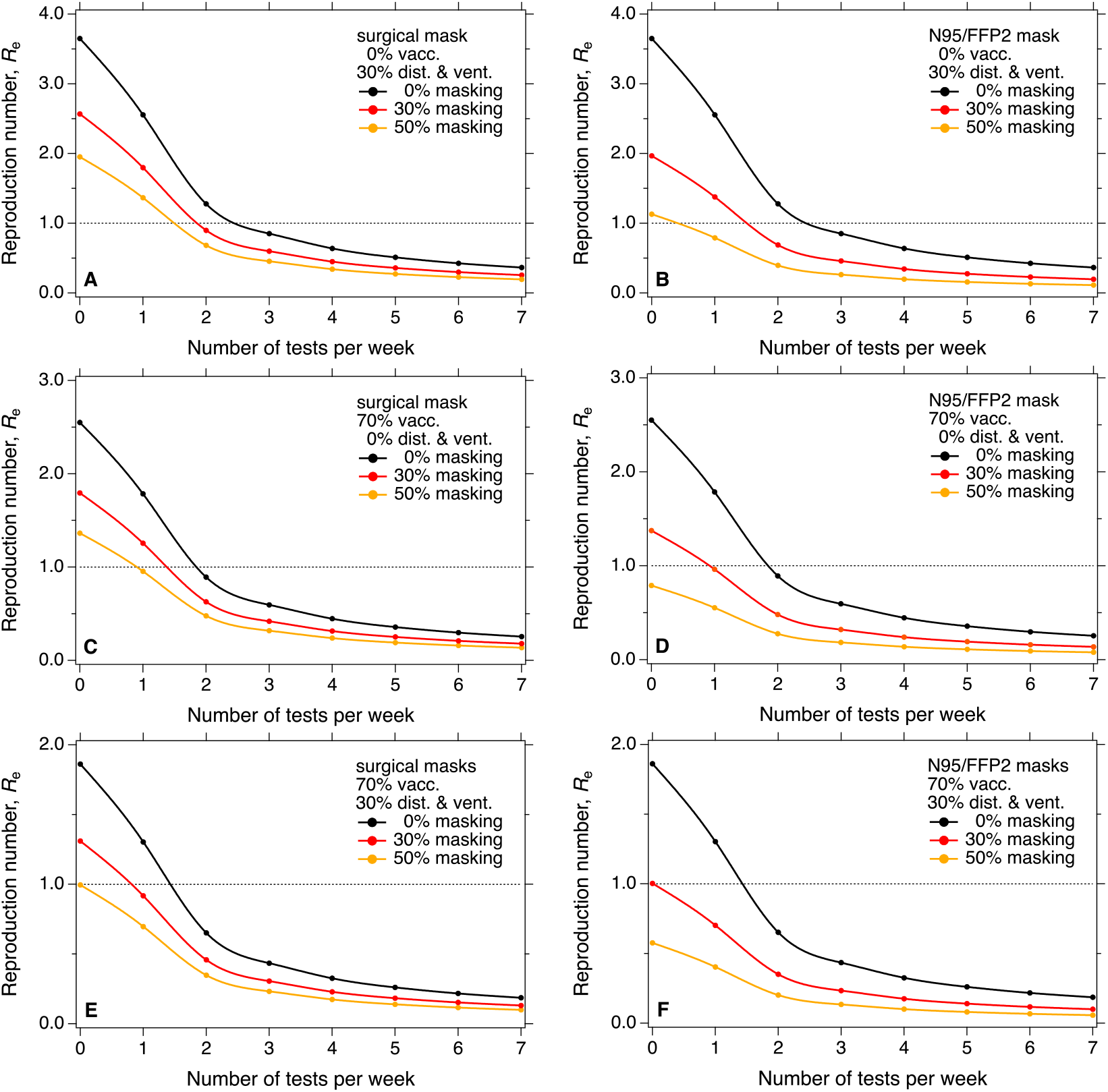
Effectiveness of testing & isolation for different compliances with masking, vaccination, and distancing & ventilation. Reduction of effective reproduction number, *R*_e_, as a function of testing frequency (number tests per week per person) for different compliances with universal masking and distancing & ventilation, as well as different vaccination rates. Basic reproduction number *R*_0_ = 5; reduced starting values of *R*_e_ reflect different vaccination rates and compliances with masking and with distancing & ventilation, respectively. Left column corresponds to universal masking with surgical masks, right column corresponds to N95/FFP2 masks.

We suggest to further extend and validate the above results by target-oriented collection and analysis of observational data. The modeling tools developed and applied in this study will be made freely available on the internet. In this context, it will be important and challenging to clarify and resolve the actual contributions of viruses in respiratory particles of different sizes, e.g., the contribution of aerosol versus droplet transmission. This will be worthwhile for both the traditional medical cut-off at 5 µm, distinguishing between fine and coarse particles, as well as for the physical upper limit at 0.1 mm, distinguishing between suspended aerosols and larger semi-ballistic particles, including droplets and dry particles (Supplementary text S6 and references therein). Among the simple physical protective measures, distancing works primarily against droplet transmission and ventilation against aerosol transmission (Methods)^3,5,6^. Surgical masks are effective and N95/FFP2 masks are highly effective against aerosol transmission, and both are even more effective against droplet transmission because of the higher filtering efficiency and low penetration rates for large droplets ^5^.

## Discussion

We suggest that the presented scientific approach, results, and tools can be used to design and communicate efficient strategies to contain the transmission of SARS-CoV-2 in different environments and to mitigate the COVID-19 pandemic. Our quantitative results are consistent with earlier recommendations ^2,31^, and the modeling tools can be used to explore and refine the synergetic effects of combining multiple protective measures as a function of *R*_0_, compliance, and efficacy of each measure (Supplementary text S6). For example, universal masking should be promoted and the efficacy and suitability of different masks against aerosol and droplet transmission under different conditions should be further clarified and communicated – in particular, why any decent mask is better than none, why tightly fitting FFP2 masks are particularly effective, and why masks are also useful in outdoor gatherings ^5,26^. Efficient ventilation of classrooms and other indoor environments could be fostered, optimized and assessed by readily available techniques like exhaust fans, air ducts for displacement ventilation, and CO_2_ sensors etc. For schools, we find that the transmission of SARS-CoV-2 can be contained by 2-3 tests per week combined with masking, distancing & ventilation, even at moderate compliances and low vaccination rates for *R*_0_ = 5. Thus, testing appears worthwhile in schools and other densely occupied environments ^14^. The frequency of testing may be adjusted according to the non-linear relation to *R*_e_ as well as the rates of false negative results ^22^, the effects of incomplete isolation, and potential changes of *R*_0_ in case of more contagious variants (see Fig. S10 for *R*_0_ = 8 to 18).

The strong dependence of *R*_e_ on practical compliance with preventive measures highlights the importance of situations where masking, distancing & ventilation or isolation are not possible, impractical, or ineffective – in particular during eating/drinking in restaurants/bars, schools/kindergartens, trains/planes, and at home. In such situations, it may, for example, help to wear masks alternatingly. Obviously, infectious fluids can also be transferred via surface contacts, and standard hygiene procedures against fomite transmission should also be followed (https://www.cdc.gov/coronavirus/2019-ncov/more/science-and-research/surface-transmission.html).

The simple and robust methods and the easy-to-understand plots of *R*_e_ vs. compliance provided in this study may help to communicate these strategies and to demonstrate the importance of cooperation to the wider public. Moreover, they may help to convince both the public and decision makers that each of the currently available measures by itself is insufficient to contain the transmissions of SARS-CoV-2 and that the synergetic effects of multiple protective measures can and have to exploited for efficient mitigation of the pandemic. Even with high and increasing rates of vaccination, other protective measures and synergetic effects should be maintained to prevent, suppress, or break potential and ongoing waves of infection. These aspects and their quantitative description might become even more important, if more transmissible variants and escape mutations of SARS-CoV-2 were to emerge. The challenge we are facing now is not just how to end the pandemic, but also how fast we can do it to save more lives and reduce the probability of further dangerous mutations of SARS-CoV-2.

## Data Availability

All data produced in the present study are available upon reasonable request to the authors once the manuscript is published in peer-review journals.

## Contributors

H.S. and Y.C. designed and led the study. Y.C., H.S. and U.P. performed the research. N.M. contributed to the calculations. C.W. contributed to the discussion. H.S., Y.C., and U.P. wrote the manuscript with input from all coauthors.

## Data monitoring committee

Hang Su, Nan Ma

## Data sharing

All data generated or analysed during this study are available in Open Research Data Repository of the Max Planck Society.

## Acknowledgments

This study is supported by the Max Planck Society (MPG), Y.C. thanks the Minerva Program of MPG.We acknowledge and emphasize the importance of Open Access to the studies and materials referenced and used in our investigations. Our research profits from Open Access policies for COVID-19-related publications, and our experience confirms that Open Access indeed accelerates scientific progress and should be extended as widely as possible.

## Supplementary Appendix

This appendix formed part of the orignal submission. We post it as supplied by the authors.

Supplement to: Su et al., Synergetic measures to contain highly transmissible variants of SARS-CoV-2

## Supplementary text

### S1. Basic reproduction number and population average infection probability

The COVID-19 pandemic has severe health, economic, and societal effects ^1^. The basic reproduction numbers, *R*_0_, for the ancestral strain of SARS-CoV-2 and Delta variant have been estimated to be ∼ 2.9 (2.4–3.4) (Billah et al., 2020)^2^ and ∼5.1 (Liu and Rocklöv, 2021)^3^, respectively. In this study, we used *R*_0_ = 3 and 5 to approximate the transmissibility of them, respectively. As the Omicron variant is suspicious to have a higher transmissibility than Delta ^4-7^, we investigated the scenarios with *R*_0_ = 8, 12 and 18. Here, *R*_0_ = 12 and 18 approximates highly contagious variants with transmissibilities similar to measles ^8^.

By definition^9^, *R*_0_, can be linked to the basic population average infection probability, *P*_0_, by *R*_0_ = *P*_0_ · *c* · *d*. Here, *d* represents the average duration of infectiousness, and *c* represents the average daily number of human contacts. As in Cheng et al. (2021), we assumed that for SARS-CoV-2 the average daily numbers of human contacts *c* ≈ 10 to 25 contacts per day and the average duration of infectiousness *d* ≈ 10 days ^10-12^. Most patients with more severe-to-critical illness or those who are severely immunocompromised likely remain infectious no longer than 20 days after symptom onset; however, there have been several reports of severely immune compromised people shedding replication-competent virus beyond 20 days” ^13-17^. Reducing the number of contacts leads to a directly proportional decrease of *R*_e_ ^9^, and as described in Method, the effects of testing & isolation of infected persons ^18^ works in a non-linear way on reducing *R*_e_.

### S2. Efficacy of vaccination against SARS-CoV-2 transmission

Based on recent observations, we assume that the probability of SARS-CoV-2 transmission is on average reduced by approx. 70% for vaccinated persons ^19-22^. In other words, the currently available vaccines are highly protective against the disease and severe outcomes of COVID-19 for both the ancestral strain of SARS-CoV-2, Delta and previous variants ^19-22^. Their efficacy against the Omicron variant is still under investigation^23^. The impact of different vaccination efficacies is illustrated in Fig. S1. Although our results show that vaccinations are currently not sufficient to contain and end the transmission of SARS-CoV-2 without synergetic measures, we are not suggesting to promote the non-pharmaceutical measures without vaccination, which would also be missing the benefit of reducing both the transmission of the virus and the severity of the disease by immunization ^19-22,24^.

### S3. Universal masking: source control, wearer protection, and practical compliance

Masks can prevent infections in two ways: (i) source control, reducing the emission and spread of respiratory viruses through airborne droplets and aerosols, and (ii) wearer protection, reducing the inhalation of airborne respiratory viruses. Surgical and N95/FFP2 masks are highly effective against aerosol transmissions ^25^, and are even more effective against droplet transmissions because of the higher filtering efficiency of masks against large droplets ^26,27^.

As explained in the main text, the fractional contribution of droplet transmission through the eyes (eye infection) is described by the parameter *x*. In Fig. S2, we show how different values of *x* would affect the results. For our calculations, we assumed an estimate of *x* = 0.3. Higher values would reduce the differences between surgical and N95/FFP2 masking.

For universal masking, 100% compliance would be difficult to achieve because masking is not always possible and practical, for example at home, during eating or drinking in restaurants and bars, in schools and kindergartens, etc. ^12,28^. The potential importance of such situations is demonstrated by a recent modeling study attributing around 10% to 40% of daily infections to restaurants and cafés/bars ^29^. Moreover, a lack of willingness to follow recommendations or mandates for mask use may also lead to low compliance with mask wearing. For example, inpatient respiratory protection studies show that adherence rates vary from 10% to 84% for health care personnel ^30,31^. Similar effects can be expected when wearing masks with low efficiency or poor fit and high penetration or leakage rates ^28,32^. Combining these effects, we may estimate ∼50% as an effective upper limit for the compliance with universal masking.

### S4. Distancing & ventilation

Physical distancing strongly reduces the transmission by droplets (> 0.1 mm), but has much smaller impact on reducing exposure to equilibrated aerosols in indoor environment ^28,33-35^. By assuming characteristic distances around ∼ 0.25 m for close contact, our calculation shows that the mass of large respiratory droplets would decrease by ∼88% at a distance of 1 m and by ∼95% at a distance of 2 m (Fig. S2). These estimates are consistent with the ∼ 60% to 90% effectiveness of physical distancing as reported in the review of Chu et al. ^36^ based on observations and data for SARS-CoV-1 and MERS ^37^. Thus, we assume approximately ∼90% (upper bound) for the effectiveness of proper physical distancing (> 1-2 m) against droplet transmission.

In contrast to physical distancing, regular ventilation has little effect on direct transmission by ballistic droplets (> 0.1 mm) but reduces indirect transmission by aerosols (< 0.1 mm) in indoor environments both in the near-field and far-field ^28,33-35,38^. Changing from passive ventilation to a high ventilation rate (e.g., 12 h^-1^, as applied in airborne infection isolation rooms) can reduce the airborne virus concentration and probability transmission with an effectiveness up to ∼90% ^28^, which can be regarded as an approximate upper bound for the effectiveness of ventilation against aerosol transmission ^38,39^. Efficient ventilation of classrooms and other indoor environments could be fostered, optimized, and assessed by readily available techniques like exhaust fans, air ducts for displacement ventilation, and CO_2_ sensors etc. ^38-40^.

The influence of distancing & ventilation on *R*_e_ is described by *E*_d&v_. The reason for combining these measures is that each of them influences only one branch of airborne transmission, either droplet transmission (distancing) or aerosol transmission (ventilation). Moreover, the relative contributions of aerosol and droplet transmission in SARS-CoV-2 transmission have not yet been determined conclusively, but they exhibit similar upper limits of effectiveness (∼90%). Thus, we assume *E*_d&v_ ≈ 90%. In summary, physical distancing by at least 1-2 meters and proper ventilation of indoor environments can decrease the risk of droplet (> 0.1 mm) and aerosol transmission (< 0.1 mm) in indoor environments by approx. 90%, whereby distancing primarily reduces droplet transmission and ventilation primarily reduces aerosol transmission ^28,38,39,41,42^. In our calculations, we assume that the population-average transmission occurs in a virus-limited regime as discussed in Cheng et al. (2021) ^28^, which would hold for the range of *R*_0_ and *R*_e_ values investigated in this study. Compared to *E*_vacc_ and *E*_mask_, the estimated value of *E*_d&v_ is subject to larger uncertainties. It should be further tested and validated, e.g., by large-scale randomized control trials, in the future.

### S5. Testing & isolation

The influence of testing & isolation on *R*_e_ is described by *E*_*t&i*_. The effects of testing & isolation are equivalent to reducing the average duration of infectiousness, *d*. According to U.S. CDC, “patients with mild-to-moderate COVID-19 remain infectious no longer than 10 days after symptom onset. Most patients with more severe-to-critical illness or those who are severely immunocompromised likely remain infectious no longer than 20 days after symptom onset; however, there have been several reports of severely immune compromised people shedding replication-competent virus beyond 20 days” ^17^. For this study, we assumed that *d* is approximately 10 days ^10^ (Supplementary text S1). When applying tests on *n* days per week, the duration of infectiousness can be effectively reduced to 7/*n* days, assuming 100% precision of tests and immediate isolation preventing further transmission. Under these conditions, we obtain *E*_*t&i*_ = (1 – 0.7/*n*). In practice, the frequency of testing has to be adjusted according to the rates of false negative results ^43^, and the effects of incomplete isolation have to be considered, which may reduce the effectiveness of this measure. The influence of a false negative testing rate *f* on *E*_*t&i*_ can be described by:

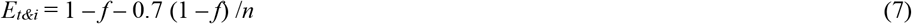

For example, a 20% false negative rate (*f* = 20%) will reduce the effectiveness of two tests per week (*n* = 2) from 65% to 52%.

### S6. Droplet versus aerosol transmission

To distinguish between aerosol transmission and droplet transmission, medical studies have frequently referred to a diameter of 5 µm as a boundary between aerosol particles and droplets, whereas in aerosol science, 100 µm or 0.1 mm are considered as an upper limit to the aerosol size range ^28,33-35,42,44-46^. Around 0.1 mm, the gravitational settling velocity of particles approaches 1 m s^-1^ in still air. Droplets larger than 0.1 mm are also called semi-ballistic or ballistic droplets, corresponding to Reynolds numbers larger than 1 or 1000, respectively^44^. They are not floating in the air but falling to the ground in seconds within a 1-2 m radius around the emitting person, approximately following a ballistic trajectory ^44^.

In the physical sciences, particles in the size range of approximately 5 to 100 µm are still considered aerosols because they remain suspended up to tens of minutes and can float many meters away from the emitter. Aerosol particles with diameters less than 5 µm can remain suspended and accumulate over hours before being removed by slow sedimentation or diffusion to the ground and walls of indoor environments.

In aerosol research and air quality monitoring, the boundary between fine and coarse aerosol particles is usually set around 2.5 µm. Fine particulate matter with diameters up to 2.5 µm (PM2.5) is known to penetrate deeper into human lungs and cause more severe health effects than coarse particulate matter ^44-48^. Actually, the air quality boundary of 2.5 µm approximately coincides with the traditional medical boundary of 5 µm, because the diameter of respiratory droplets emitted at 100% relative humidity decreases by about one half upon drying in ambient air at typical relative humidities in the range of 30-60%, e.g., from 5 µm to approx. 2.5 µm ^25^. In analogy to atmospheric PM2.5, fine respiratory aerosol particles may also penetrate deeper into the human lungs, and we hypothesize that this might cause more severe infections, which remains to be elucidated.

With regard to the relevance and occurrence of aerosol vs. droplet transmission, the following relations are relevant regardless of the boundary used to distinguish between aerosols and larger droplets (5 µm or 0.1 mm). If the viral load (concentration) in epithelial lining fluids and in respiratory aerosol particles or droplets is small, or if the median infectious dose of the virus is large, then the total volume of respiratory aerosol particles (<5 µm or <0.1 mm) and the virus dose contained in aerosols may be too small to cause substantial infection risks by aerosol transmission. Under such circumstances, only droplets in the larger size ranges (>5 µm or > 0.1 mm) with a large total volume are expected to contribute substantially to the infection probability and virus transmission (“droplet transmission”).

Even if the viral load (concentration) in large respiratory droplets may be lower than in small respiratory aerosols due to different origins and enrichment processes in the respiratory tract ^25^, the size dependence of the particle volume is so strong (diameter to the power of three) that it seems unlikely that virus transmission would proceed only through aerosols and not through larger droplets as well. On the contrary, it seems that aerosol and droplet transmission are likely to go hand in hand. For direct infections in the near-field, droplet transmission is likely to be even more important than aerosol transmission, unless there is an enormous size dependence of viral load (more than three orders of magnitude). Therefore, masking and distancing are important precautions during contacts also outdoors, not just in indoor environments. In practice, the relative importance of aerosol and droplet transmission has not yet been measured and resolved for SARS-CoV-2, regardless of the definition and boundary applied (5 µm or 0.1 mm).

By consistent use of masks, distancing & ventilation as well as testing & isolation, it was possible to contain early variants of SARS-CoV-2 even without vaccination as demonstrated in several Asia-Pacific countries during the years 2020-2021^49-52^. It will depend on the basic reproduction number of Omicron and further variants whether these measures may suffice or more strict measures like sustained or recurring general contact reductions may be required to mitigate the pandemic and prevent or control future waves of infection.

We suggest that the presented scientific approach, results, and tools can be used to design and communicate efficient strategies to contain the transmission of SARS-CoV-2 in different environments and to mitigate the COVID-19 pandemic. Our quantitative results are consistent with earlier recommendations ^53-55^, and the modeling tools can be used to explore and refine the synergetic effects of combining multiple protective measures as a function of *R*_0_, compliance, and efficacy of each measure.

**Fig. S1.**
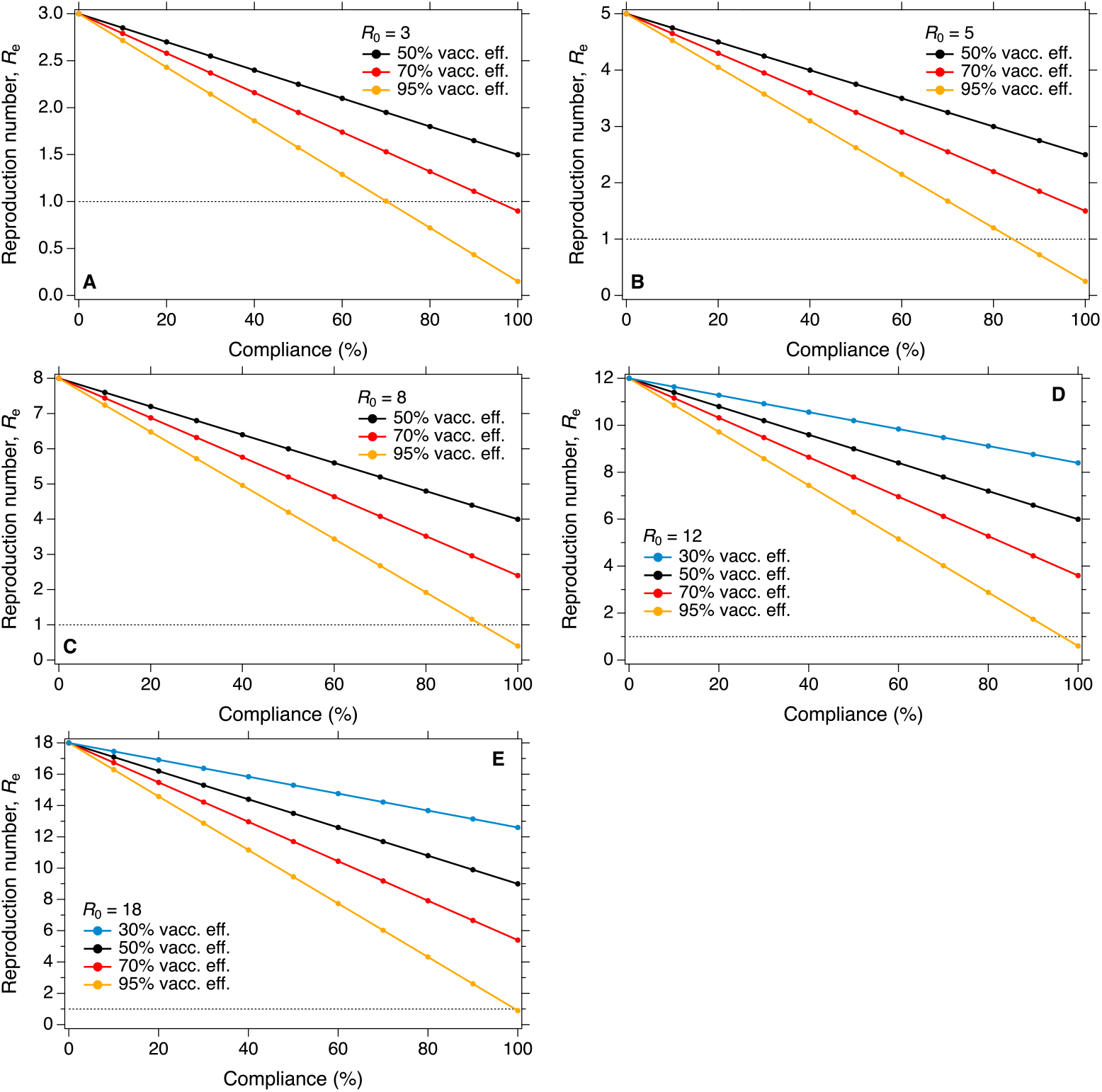
Effectiveness of vaccination for different vaccination rates and vaccination efficiency. Reduction of effective reproduction number, *R*_e_, as a function of vaccination efficiency (“vacc. eff.”) for different vaccination rates (“compliance” in %). Panel **A** is for a basic reproduction number *R*_0_ = 3 approximately reflecting the transmissibility of the ancestral strain of SARS-CoV-2. Panel **B** is for *R*_0_ = 5 approximately reflecting the transmissibility of the delta variant of SARS-CoV-2. Panel **C** is for *R*_0_ = 8 approximating a variant with higher transmissibility than the delta variant of SARS-CoV-2. Panel **D** is for *R*_0_ = 12 and panel **E** is for *R*_0_ = 18 approximating the measles-like transmissibility.

**Fig. S2.**
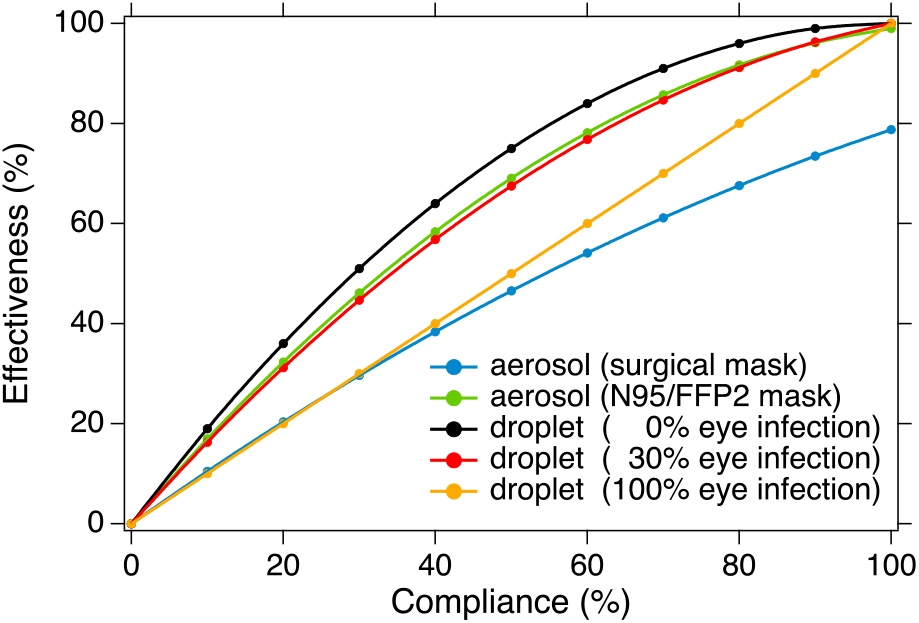
Effectiveness of masks in preventing aerosol or droplet transmission. The blue and green lines represent the effectiveness of surgical masks and N95/FFP2 masks in preventing aerosol transmission, respectively. The rest lines represent the effectiveness of masks in preventing droplet transmission with different contributions of eye infections (See Method).

**Fig. S3.**
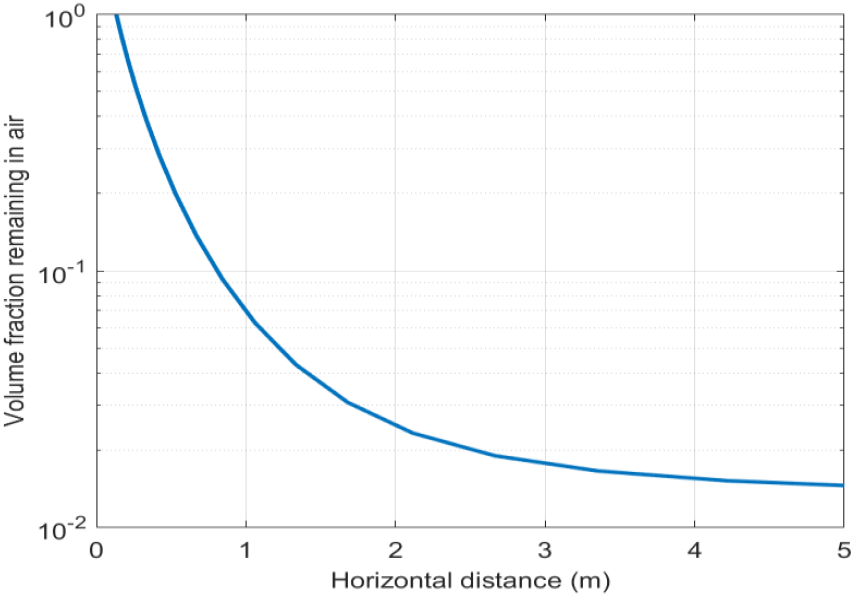
Fraction of remaining respiratory particle volume (aerosols and droplets with diameter up to 1 mm). Assumption: after ejection from the mouth, all respiratory particles move forward at a horizontal speed of 5 m s^-1^ with the airflow and the horizontal velocity remains constant; the deposition velocity is caused by the gravitational settling and is size dependent. When the vertical settling distance of the particles is < 1.2 m, we considered them as the remaining particles that may infect others.

**Fig. S4.**
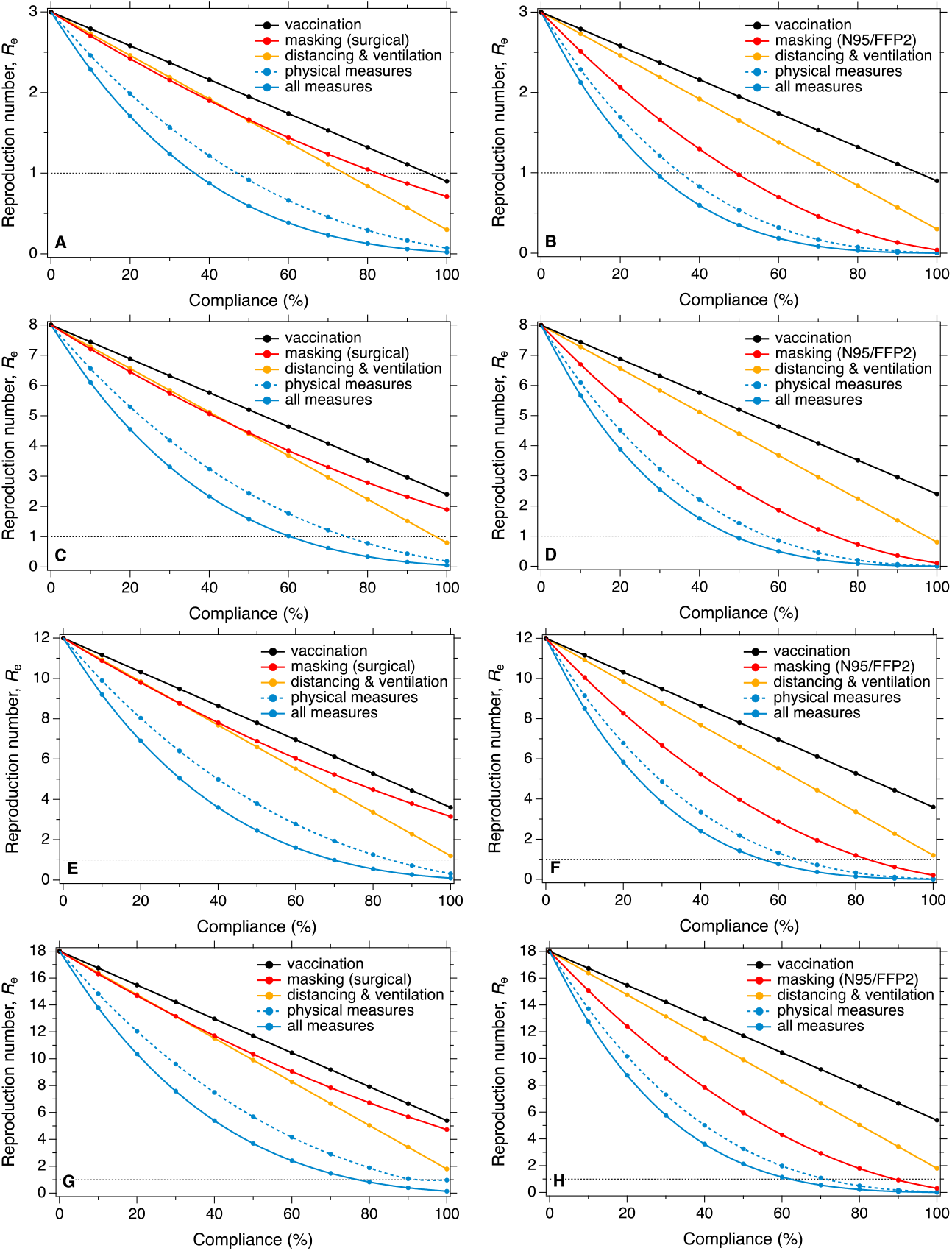
Effectiveness of individual and combined measures. Same as Fig. 2, for *R*_0_ = 3, 8, 12 and 18. Reduction of effective reproduction number, *R*_e_, as a function of compliance with different protective measures for a basic reproduction number *R*_0_ = 8 (**C** and **D**) approximating a variant with higher transmissibility than the delta variant of SARS-CoV-2 and for *R*_0_ = 3 (**A** and **B**) approximately reflecting the transmissibility of the ancestral strain of SARS-CoV-2, and for *R*_0_ = 12 (**E** and **F**) and *R*_0_ = 18 (**G** and **F**) approximating a variant with similar transmissibility of measles. The curve labeled “physical measures” refers to the combination and synergy of universal masking plus distancing and ventilation; the curve labeled “all measures” refers to the combination and synergy of the physical measures with vaccination. Left column corresponds to universal masking with surgical masks, right column corresponds to N95/FFP2 masks.

**Fig. S5.**
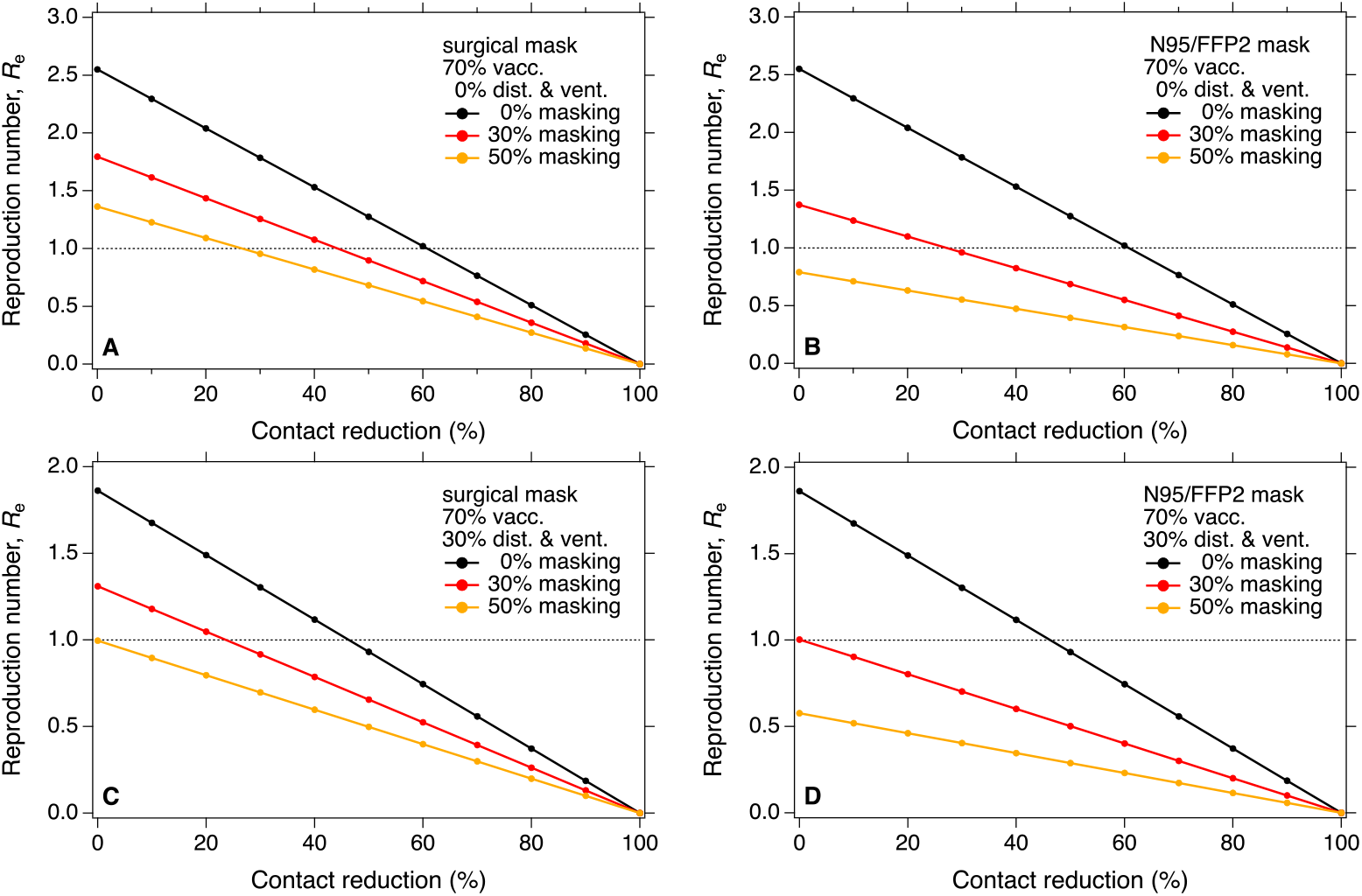
Effectiveness of contact reduction for different compliances with masking and distancing & ventilation. Reduction of effective reproduction number, *R*_e_, as a function of contact reduction for different compliances with universal masking at a fixed vaccination rate of 70%. Basic reproduction number *R*_0_ = 5; reduced starting values of *R*_e_ at zero contact reduction reflect the vaccination rate and different levels of masking and distancing & ventilation (30% compliance, panels **C** and **D**). Left column corresponds to universal masking with surgical masks, right column corresponds to N95/FFP2 masks.

**Fig. S6.**
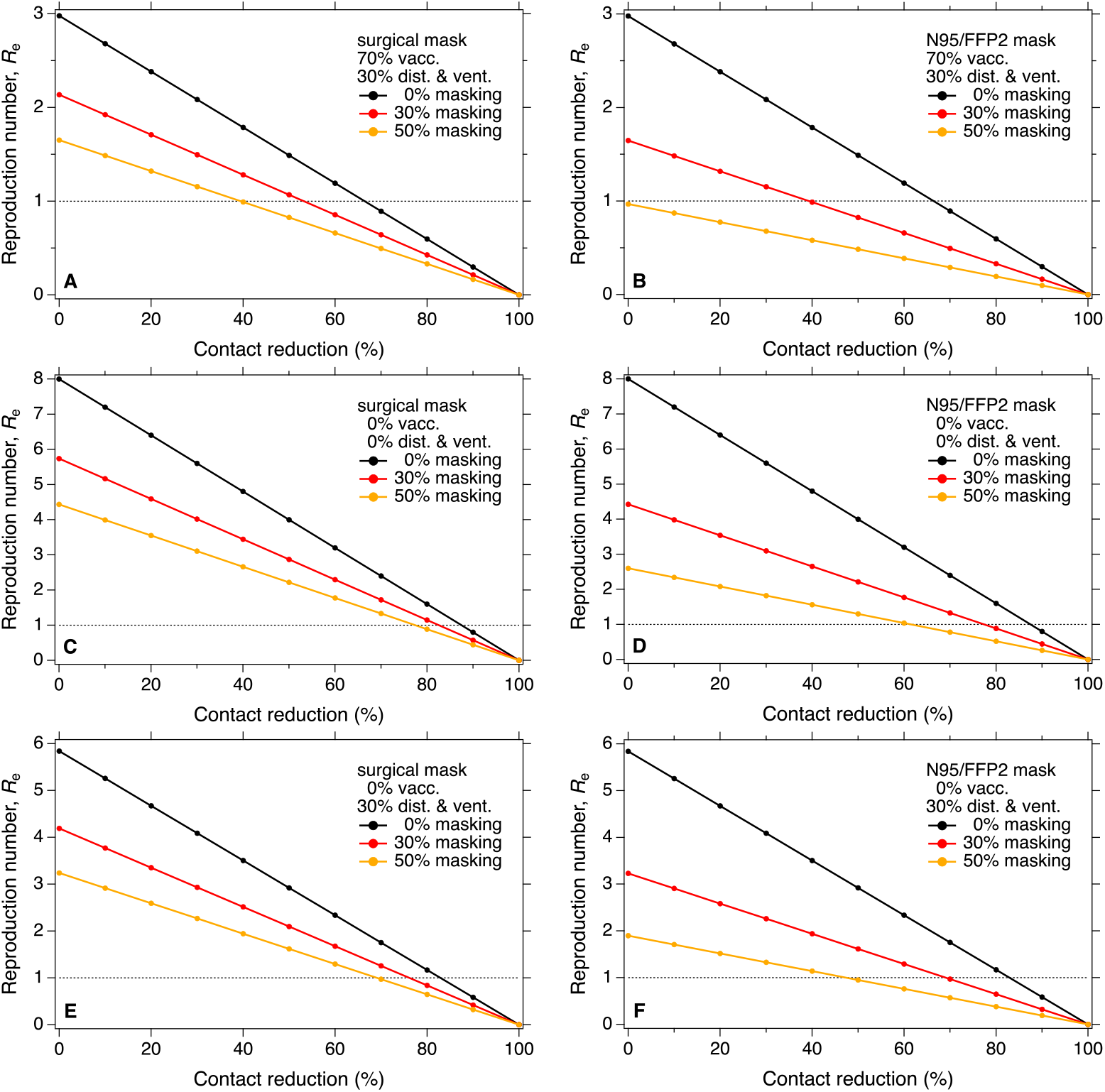
Effectiveness of contact reduction for different compliances with masking and distancing & ventilation. Similar to Fig. S5, for *R*_0_ = 8: reduction of effective reproduction number, *R*_e_, as a function of contact reduction for different compliances with universal masking at a fixed vaccination rate of 70% and 30% compliance of distancing and ventilation (**A** and **B**). In the lower four panels (**C, D, E** and **F**), we consider the worst scenario when vaccination does not work for a new highly contagious variant (0% vaccination rate). Basic reproduction number *R*_0_ = 8; reduced starting values of *R*_e_ at zero contact reduction reflect the vaccination rate and different levels of masking and distancing & ventilation. Left column corresponds to universal masking with surgical masks, right column corresponds to N95/FFP2 masks.

**Fig. S7.**
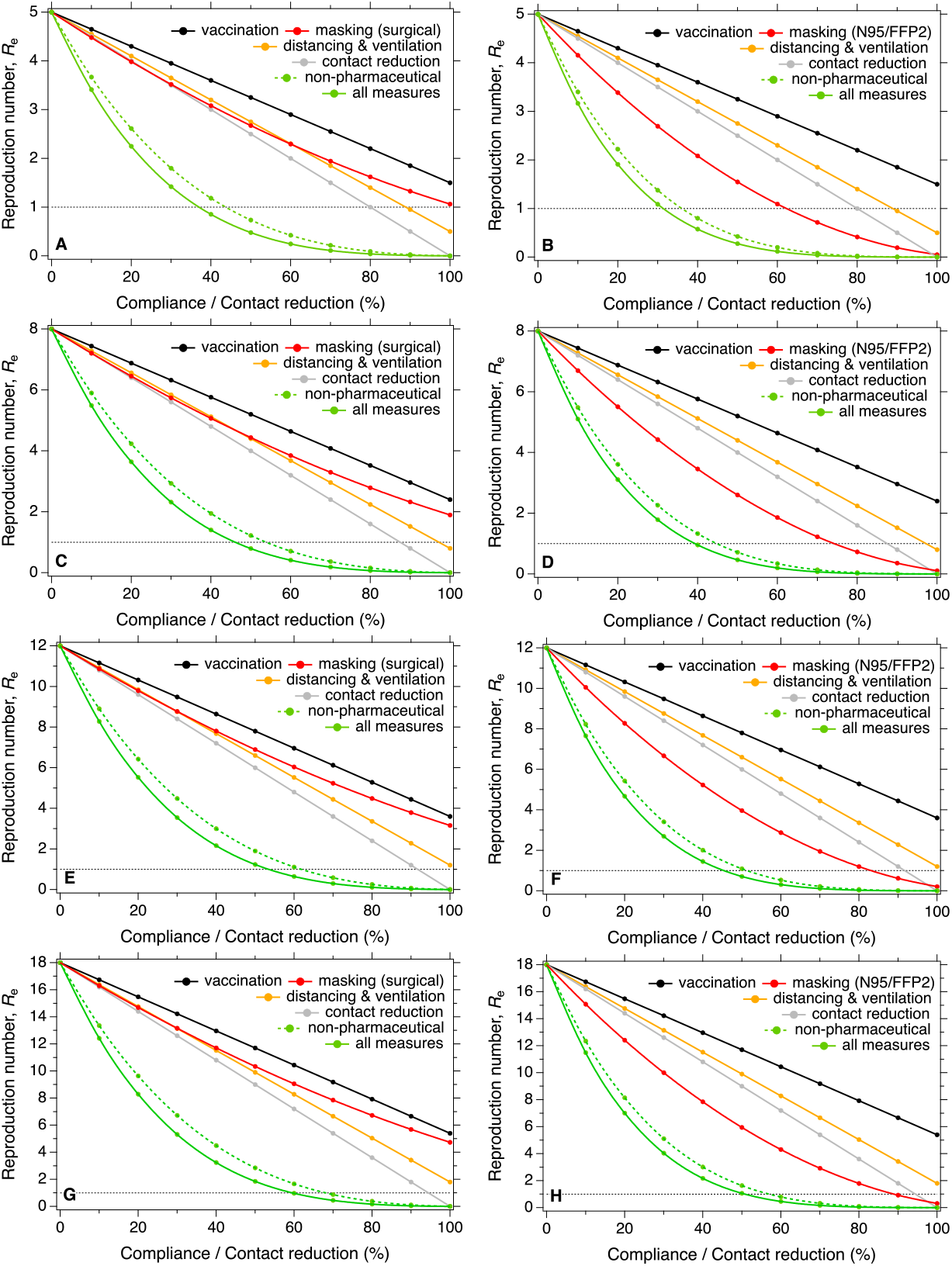
Effectiveness of individual and combined measures. Similar to Fig. 2, with contact reduction as an additional measure for *R*_0_ = 5, 8, 12 and 18. Reduction of effective reproduction number, *R*_e_, as a function of compliance with different protective measures and contact reduction for a basic reproduction number *R*_0_ = 5 (**A** and **B**) approximately reflecting the transmissibility of the delta variant of SARS-CoV-2, for *R*_0_ = 8 (**C** and **D**) approximating a variant with higher transmissibility than the delta variant, and for *R*_0_ = 12 (**E** and **F**) and *R*_0_ = 18 (**G** and **F**) approximating a variant with similar transmissibility of measles. The curve labeled “non-pharmaceutical” refers to the combination and synergy of universal masking, distancing and ventilation and contact reduction; the curve labeled “all measures” refers to the combination and synergy of the non-pharmaceutical measures with vaccination. Left column corresponds to universal masking with surgical masks, right column corresponds to N95/FFP2 masks.

**Fig. S8.**
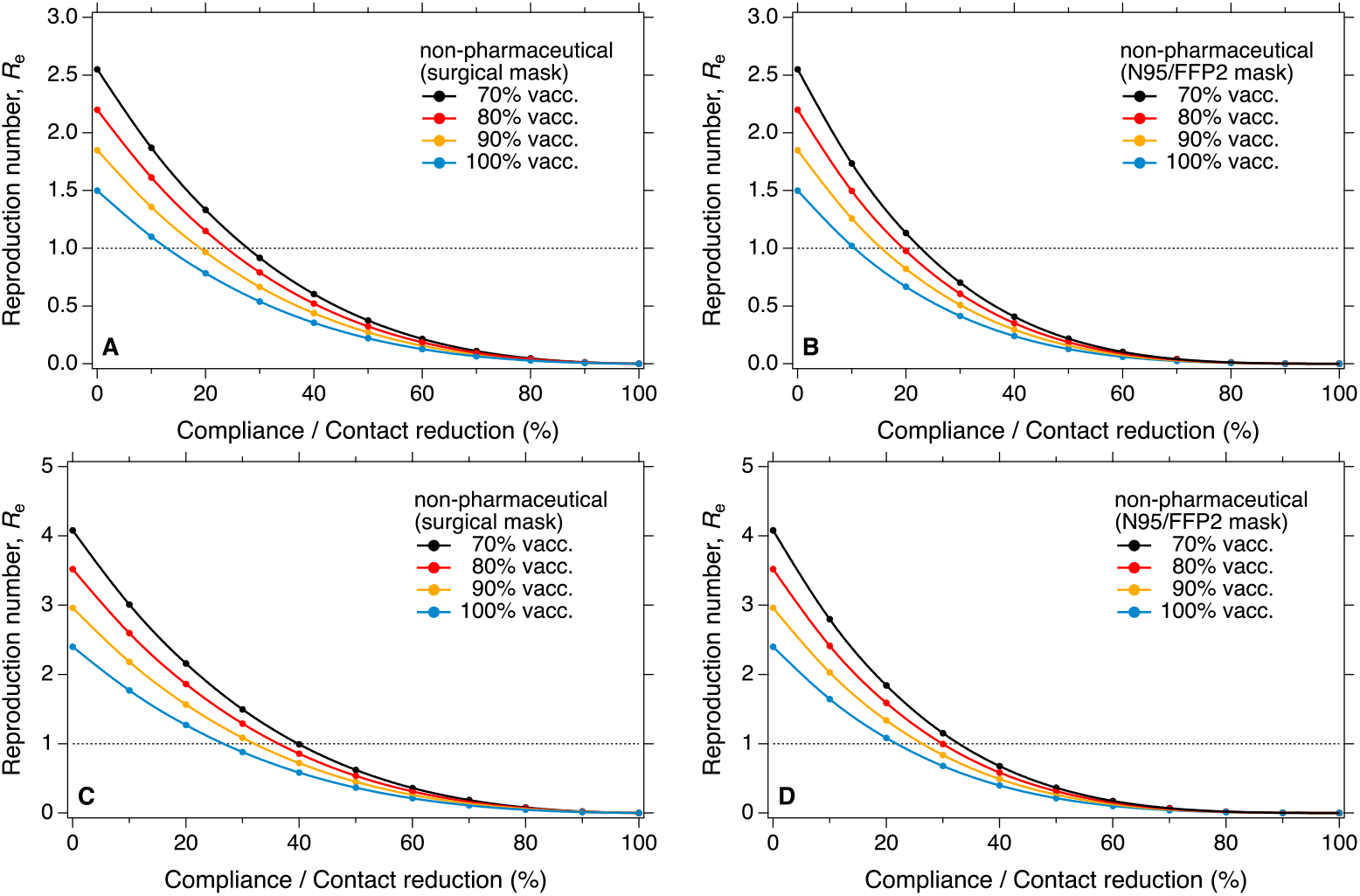
Effectiveness of combined non-pharmaceutical measures for different vaccination rates. Reduction of effective reproduction number, *R*_e_, as a function of compliance with all investigated non-pharmaceutical measures for different vaccination rates in the population. Non-pharmaceutical measures are the combination and synergy of universal masking, distancing and ventilation and contact reduction. Basic reproduction number *R*_0_ = 5 (**A** and **B**) and *R*_0_ = 8 (**C** and **D**); reduced starting values of *R*_e_ at zero compliance with non-pharmaceutical measures correspond to different vaccination rates. Left column corresponds to universal masking with surgical masks, right column corresponds to N95/FFP2 masks.

**Fig. S9.**
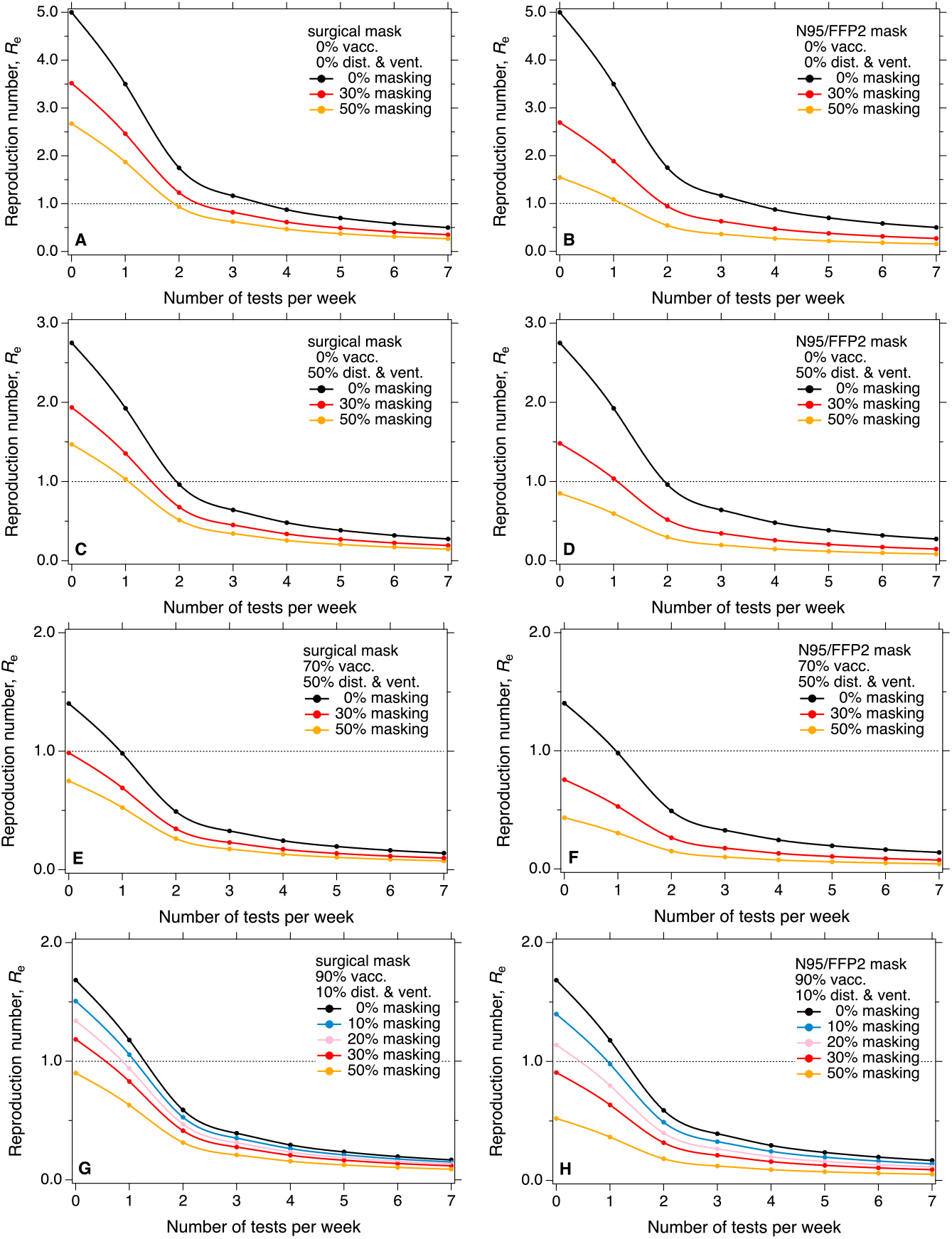
Effectiveness of testing & isolation for different compliances with masking, vaccination, and distancing & ventilation. Similar to Fig. 4, but with varies of combination of vaccination rates, compliance of universal masking and distancing & ventilation. Reduction of effective reproduction number, *R*_e_, as a function of testing frequency (number tests per week per person) for different compliances with universal masking and distancing & ventilation, as well as different vaccination rates. Basic reproduction number *R*_0_ = 5; reduced starting values of *R*_e_ reflect different vaccination rates and compliances with masking and with distancing & ventilation, respectively. Left column corresponds to universal masking with surgical masks, right column corresponds to N95/FFP2 masks.

**Fig. S10.**
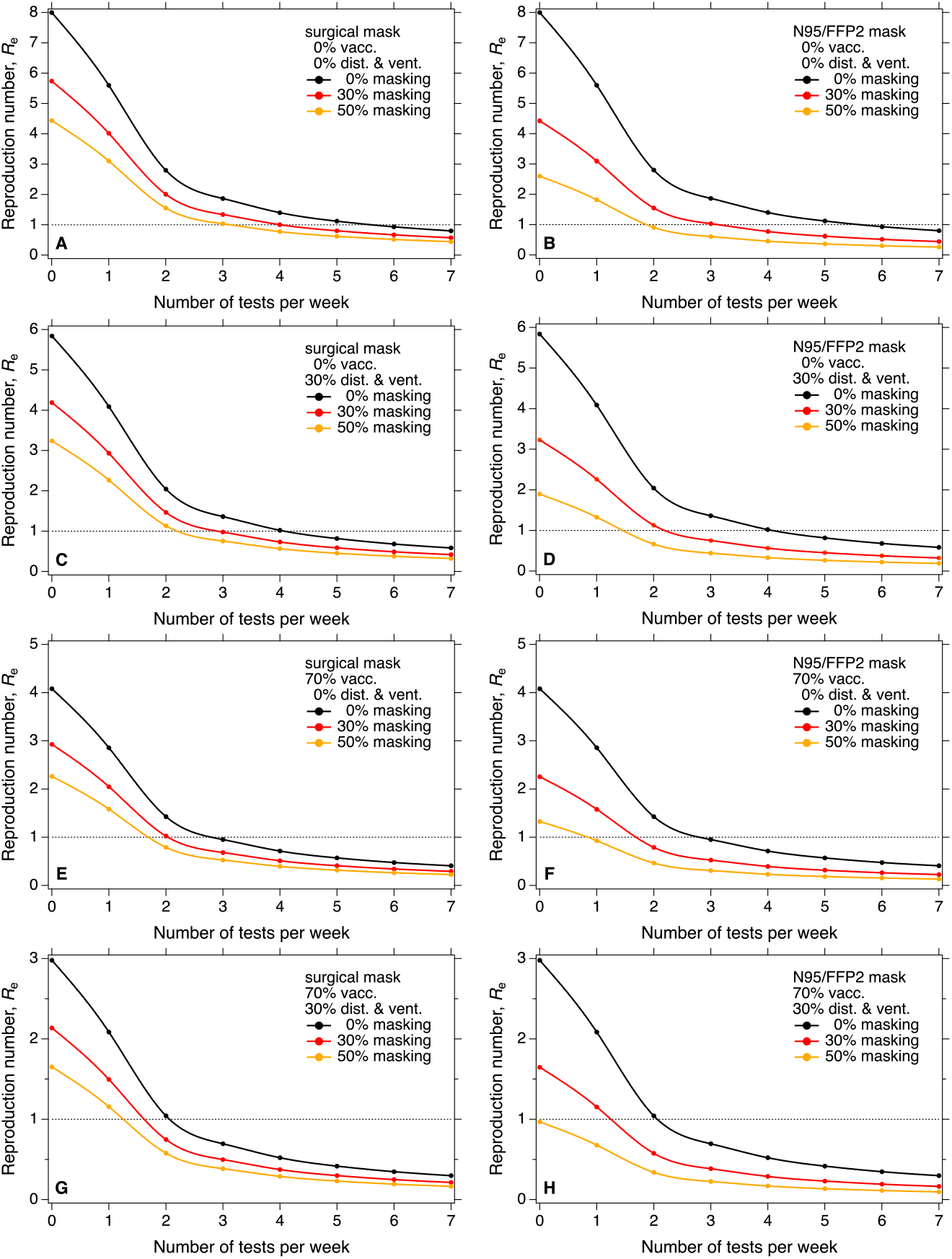
Effectiveness of testing & isolation for different compliances with masking, vaccination, and distancing & ventilation. Similar to Fig. S9, for *R*_0_ = 8. Reduction of effective reproduction number, *R*_e_, as a function of testing frequency (number tests per week per person) for different compliances with universal masking, distancing & ventilation, and contact reduction as well as different vaccination rates. Basic reproduction number *R*_0_ = 8; reduced starting values of *R*_e_ reflect different vaccination rates and compliances with masking, distancing & ventilation and contact reduction, respectively. Left column corresponds to universal masking with surgical masks, right column corresponds to N95/FFP2 masks.

**Fig. S11.**
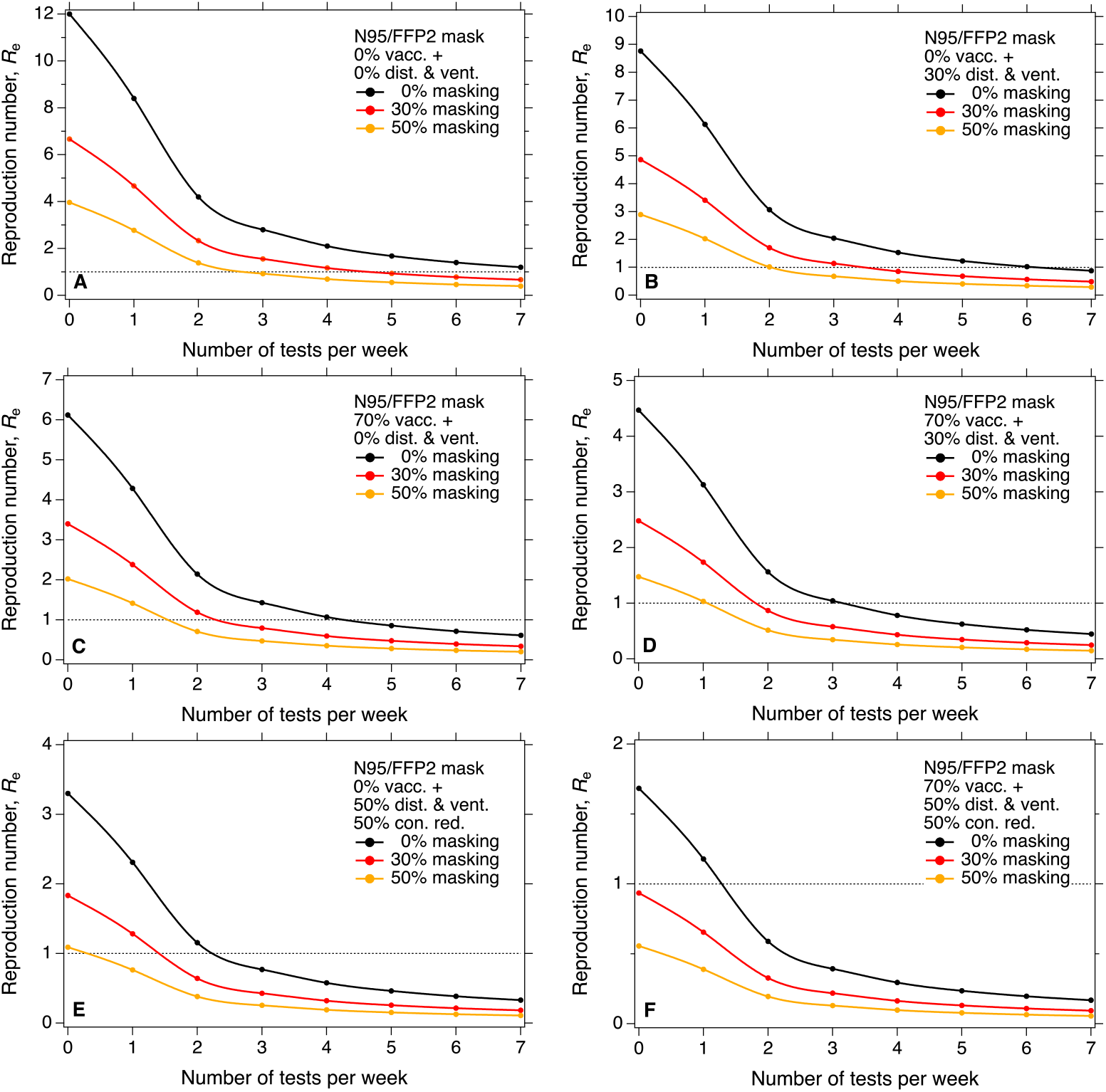
Effectiveness of testing & isolation for different compliances with masking, vaccination, distancing & ventilation, and contact reduction. Similar to Fig. S9, for *R*_0_ = 12. Reduction of effective reproduction number, *R*_e_, as a function of testing frequency (number tests per week per person) for different compliances with universal masking, distancing & ventilation, and contact reduction as well as different vaccination rates. Basic reproduction number *R*_0_ = 12; reduced starting values of *R*_e_ reflect different vaccination rates and compliances with masking, distancing & ventilation and contact reduction, respectively. Only N95/FFP2 masks are applied.

**Fig. S12.**
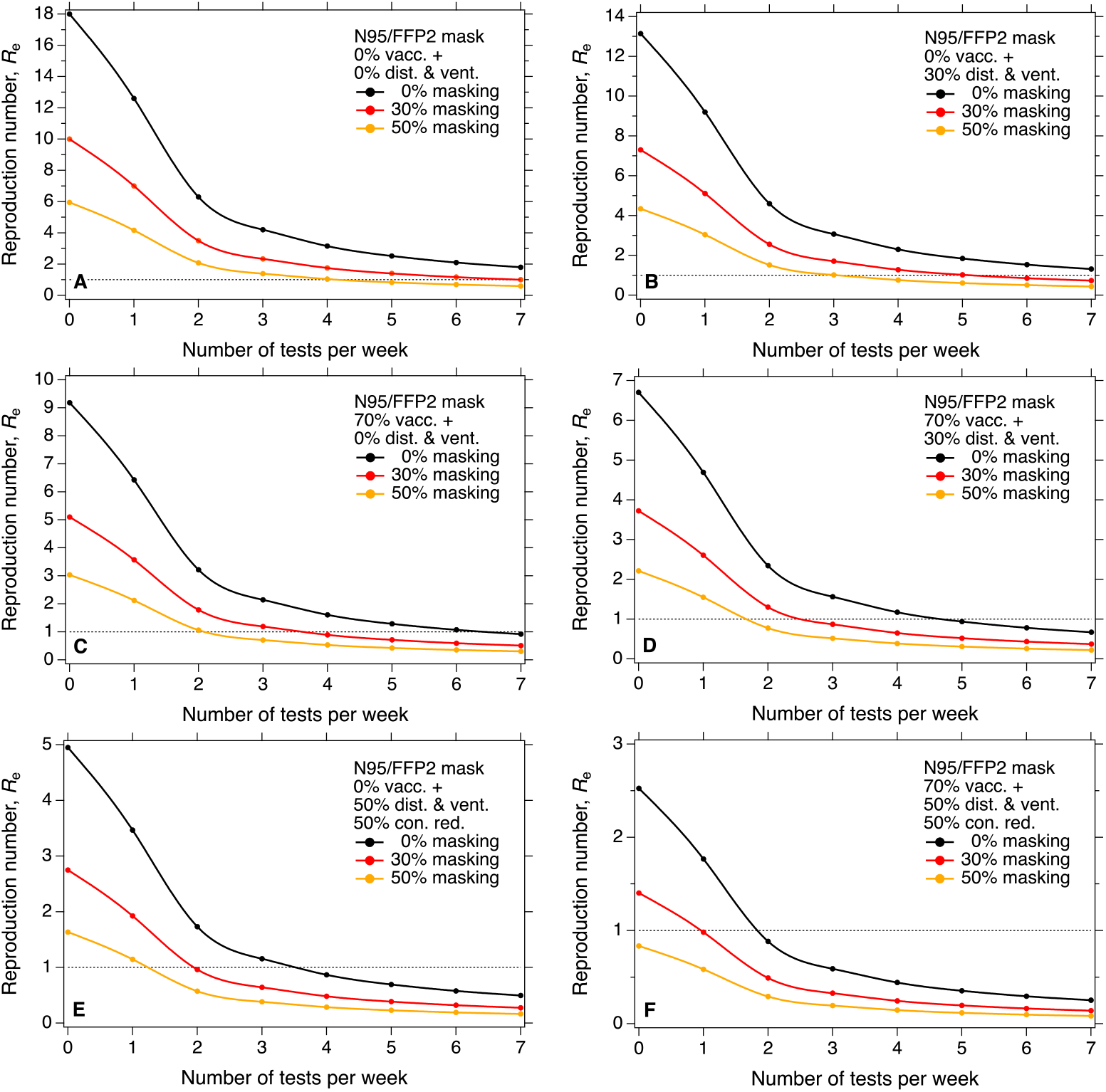
Effectiveness of testing & isolation for different compliances with masking, vaccination, distancing & ventilation, and contact reduction. Similar to Fig. S9, for *R*_0_ = 18. Reduction of effective reproduction number, *R*_e_, as a function of testing frequency (number tests per week per person) for different compliances with universal masking, distancing & ventilation, and contact reduction as well as different vaccination rates. Basic reproduction number *R*_0_ = 18; reduced starting values of *R*_e_ reflect different vaccination rates and compliances with masking, distancing & ventilation and contact reduction, respectively. Only N95/FFP2 masks are applied.

## References

1. Polack FP, Thomas SJ, Kitchin N, et al. Safety and Efficacy of the BNT162b2 mRNA Covid-19 Vaccine. New England Journal of Medicine 2020; 383(27): 2603–15.

2. Moore S, Hill EM, Tildesley MJ, Dyson L, Keeling MJ. Vaccination and non-pharmaceutical interventions for COVID-19: a mathematical modelling study. The Lancet Infectious Diseases 2021; 21(6): 793–802.

3. Wang CC, Prather KA, Sznitman J, et al. Airborne transmission of respiratory viruses. Science 2021; 373(6558): eabd9149.

4. Lelieveld J, Helleis F, Borrmann S, et al. Model Calculations of Aerosol Transmission and Infection Risk of COVID-19 in Indoor Environments. International Journal of Environmental Research and Public Health 2020; 17(21): 8114.

5. Cheng Y, Ma N, Witt C, et al. Face masks effectively limit the probability of SARS-CoV-2 transmission. Science 2021: eabg6296.

6. Morawska L, Allen J, Bahnfleth W, et al. A paradigm shift to combat indoor respiratory infection. Science 2021; 372(6543): 689–91.

7. Jones NR, Qureshi ZU, Temple RJ, Larwood JPJ, Greenhalgh T, Bourouiba L. Two metres or one: what is the evidence for physical distancing in covid-19? BMJ 2020; 370: m3223.

8. Islam N, Sharp SJ, Chowell G, et al. Physical distancing interventions and incidence of coronavirus disease 2019: natural experiment in 149 countries. BMJ 2020; 370: m2743.

9. Haug N, Geyrhofer L, Londei A, et al. Ranking the effectiveness of worldwide COVID-19 government interventions. Nature Human Behaviour 2020; 4(12): 1303–12.

10. Jason Abaluck LHK, Ashley Styczynski4, Ashraful Haque, Md. Alamgir Kabir, Ellen Bates-Jefferys, Emily Crawford, Jade Benjamin-Chung, Shabib Raihan, Shadman Rahman, Salim Benhachmi, Neeti Zaman, Peter J. Winch, Maqsud Hossain, Hasan Mahmud Reza, Abdullah All Jaber, Shawkee Gulshan Momen, Aura Rahman, Faika Laz Banti, Tahrima Saiha Huq,, Stephen P. Luby AMM. The Impact of Community Masking on COVID-19: A Cluster-Randomized Trial in Bangladesh. 2021.

11. Liu Y, Rocklöv J. The reproductive number of the Delta variant of SARS-CoV-2 is far higher compared to the ancestral SARS-CoV-2 virus. Journal of Travel Medicine 2021; 28(7).

12. Tartof SY, Slezak JM, Fischer H, et al. Effectiveness of mRNA BNT162b2 COVID-19 vaccine up to 6 months in a large integrated health system in the USA: a retrospective cohort study. The Lancet 2021; 398(10309): 1407–16.

13. Cohn BA, Cirillo PM, Murphy CC, Krigbaum NY, Wallace AW. SARS-CoV-2 vaccine protection and deaths among US veterans during 2021. Science; 0(0): eabm0620.

14. Willeit P, Krause R, Lamprecht B, et al. Prevalence of RT-qPCR-detected SARS-CoV-2 infection at schools: First results from the Austrian School-SARS-CoV-2 prospective cohort study. The Lancet Regional Health - Europe 2021; 5: 100086.

15. Bagheri G, Thiede B, Hejazi B, Schlenczek O, Bodenschatz E. An upper bound on one-to-one exposure to infectious human respiratory particles. Proc Natl Acad Sci 2021; 118(49): e2110117118.

16. Tang JW, Bahnfleth WP, Bluyssen PM, et al. Dismantling myths on the airborne transmission of severe acute respiratory syndrome coronavirus-2 (SARS-CoV-2). J Hosp Infect 2021; 110: 89–96.

17. Coroneo MT, Collignon PJ. SARS-CoV-2: eye protection might be the missing key. The Lancet Microbe 2021; 2(5): e173–e4.

18. Prather KA, Marr LC, Schooley RT, McDiarmid MA, Wilson ME, Milton DK. Airborne transmission of SARS-CoV-2. Science 2020; 370(6514): 303–4.

19. Chu DK, Akl EA, Duda S, et al. Physical distancing, face masks, and eye protection to prevent person-to-person transmission of SARS-CoV-2 and COVID-19: a systematic review and meta-analysis. The Lancet 2020; 395(10242): 1973–87.

20. Choi B, Choudhary MC, Regan J, et al. Persistence and Evolution of SARS-CoV-2 in an Immunocompromised Host. New England Journal of Medicine 2020; 383(23): 2291–3.

21. Johansson MA, Quandelacy TM, Kada S, et al. SARS-CoV-2 Transmission From People Without COVID-19 Symptoms. JAMA Network Open 2021; 4(1): e2035057–e.

22. Albert E, Torres I, Bueno F, et al. Field evaluation of a rapid antigen test (Panbio™ COVID-19 Ag Rapid Test Device) for COVID-19 diagnosis in primary healthcare centres. Clinical Microbiology and Infection 2021; 27(3): 472.e7-.e10.

23. Roser M. Coronavirus Pandemic (COVID-19). Our World in Data 2020.

24. Chang S, Pierson E, Koh PW, et al. Mobility network models of COVID-19 explain inequities and inform reopening. Nature 2021; 589(7840): 82–7.

25. Radonovich LJ, J., Simberkoff MS, Bessesen MT, et al. N95 Respirators vs Medical Masks for Preventing Influenza Among Health Care Personnel: A Randomized Clinical Trial. Jama 2019; 322(9): 824–33.

26. Howard J, Huang A, Li Z, et al. An evidence review of face masks against COVID-19. Proc Natl Acad Sci 2021; 118(4): e2014564118.

27. Aguilera Benito P, Piña Ramírez C, Viccione G, Lepore E. Ventilation for Residential Buildings: Critical Assessment of Standard Requirements in the COVID-19 Pandemic Context. Frontiers in Built Environment 2021; 7(113).

28. Flaxman S, Mishra S, Gandy A, et al. Estimating the effects of non-pharmaceutical interventions on COVID-19 in Europe. Nature 2020; 584(7820): 257–61.

29. Pullano G, Valdano E, Scarpa N, Rubrichi S, Colizza V. Evaluating the effect of demographic factors, socioeconomic factors, and risk aversion on mobility during the COVID-19 epidemic in France under lockdown: a population-based study. The Lancet Digital Health 2020; 2(12): e638–e49.

30. Schlosser F, Maier BF, Jack O, Hinrichs D, Zachariae A, Brockmann D. COVID-19 lockdown induces disease-mitigating structural changes in mobility networks. Proc Natl Acad Sci 2020; 117(52): 32883–90.

31. Dehning J, Zierenberg J, Spitzner FP, et al. Inferring change points in the spread of COVID-19 reveals the effectiveness of interventions. Science 2020; 369(6500): eabb9789.

## References

1. Castillo JC, Ahuja A, Athey S, et al. Market design to accelerate COVID-19 vaccine supply. Science 2021; 371(6534): 1107–9.

2. Billah MA, Miah MM, Khan MN. Reproductive number of coronavirus: A systematic review and meta-analysis based on global level evidence. PLoS One 2020; 15(11): e0242128.

3. Liu Y, Rocklöv J. The reproductive number of the Delta variant of SARS-CoV-2 is far higher compared to the ancestral SARS-CoV-2 virus. Journal of Travel Medicine 2021; 28(7).

4. WHO. Enhancing Readiness for Omicron (B.1.1.529): Technical Brief and Priority Actions for Member States, 2021.

5. Neil Ferguson AG, Anne Cori, Alexandra Hogan, Wes Hinsley, Erik Volz. Report 49 - Growth, population distribution and immune escape of Omicron in England, 2021.

6. Control ECfDPa. Implications of the further emergence and spread of the SARS-CoV-2 B.1.1.529 variant of concern (Omicron) for the EU/EEA – first update, 2021.

7. Omicron Variant: What You Need to Know. 2021. https://www.cdc.gov/coronavirus/2019-ncov/variants/omicron-variant.html.

8. Guerra FM, Bolotin S, Lim G, et al. The basic reproduction number (R(0)) of measles: a systematic review. Lancet Infect Dis 2017; 17(12): e420–e8.

9. van den Driessche P, Watmough J. Further Notes on the Basic Reproduction Number. In: Brauer F, van den Driessche P, Wu J, eds. Mathematical Epidemiology. Berlin, Heidelberg: Springer Berlin Heidelberg; 2008: 159–78.

10. Johansson MA, Quandelacy TM, Kada S, et al. SARS-CoV-2 Transmission From People Without COVID-19 Symptoms. JAMA Network Open 2021; 4(1): e2035057–e.

11. Zhang J, Litvinova M, Liang Y, et al. Changes in contact patterns shape the dynamics of the COVID-19 outbreak in China. Science 2020; 368(6498): 1481–6.

12. Mossong J, Hens N, Jit M, et al. Social Contacts and Mixing Patterns Relevant to the Spread of Infectious Diseases. PLOS Medicine 2008; 5(3): e74.

13. Aydillo T, Gonzalez-Reiche AS, Aslam S, et al. Shedding of Viable SARS-CoV-2 after Immunosuppressive Therapy for Cancer. New England Journal of Medicine 2020; 383(26): 2586–8.

14. Avanzato VA, Matson MJ, Seifert SN, et al. Case Study: Prolonged Infectious SARS-CoV-2 Shedding from an Asymptomatic Immunocompromised Individual with Cancer. Cell 2020; 183(7): 1901–12.e9.

15. Baang JH, Smith C, Mirabelli C, et al. Prolonged Severe Acute Respiratory Syndrome Coronavirus 2 Replication in an Immunocompromised Patient. The Journal of Infectious Diseases 2020; 223(1): 23–7.

16. Tarhini H, Recoing A, Bridier-nahmias A, et al. Long-Term Severe Acute Respiratory Syndrome Coronavirus 2 (SARS-CoV-2) Infectiousness Among Three Immunocompromised Patients: From Prolonged Viral Shedding to SARS-CoV-2 Superinfection. The Journal of Infectious Diseases 2021; 223(9): 1522–7.

17. Choi B, Choudhary MC, Regan J, et al. Persistence and Evolution of SARS-CoV-2 in an Immunocompromised Host. New England Journal of Medicine 2020; 383(23): 2291–3.

18. Willeit P, Krause R, Lamprecht B, et al. Prevalence of RT-qPCR-detected SARS-CoV-2 infection at schools: First results from the Austrian School-SARS-CoV-2 prospective cohort study. The Lancet Regional Health - Europe 2021; 5: 100086.

19. Tartof SY, Slezak JM, Fischer H, et al. Effectiveness of mRNA BNT162b2 COVID-19 vaccine up to 6 months in a large integrated health system in the USA: a retrospective cohort study. The Lancet 2021; 398(10309): 1407–16.

20. Tang P, Hasan MR, Chemaitelly H, et al. BNT162b2 and mRNA-1273 COVID-19 vaccine effectiveness against the SARS-CoV-2 Delta variant in Qatar. Nature Medicine 2021.

21. Fowlkes A, Gaglani M, Groover K, et al. Effectiveness of COVID-19 Vaccines in Preventing SARS-CoV-2 Infection Among Frontline Workers Before and During B.1.617.2 (Delta) Variant Predominance - Eight U.S. Locations, December 2020-August 2021. MMWR Morb Mortal Wkly Rep 2021; 70(34): 1167–9.

22. Cohn BA, Cirillo PM, Murphy CC, Krigbaum NY, Wallace AW. SARS-CoV-2 vaccine protection and deaths among US veterans during 2021. Science; 0(0): eabm0620.

23. Andrews N, Stowe J, Kirsebom F, et al. Effectiveness of COVID-19 vaccines against the Omicron (B.1.1.529) variant of concern. medRxiv 2021: 2021.12.14.21267615.

24. Polack FP, Thomas SJ, Kitchin N, et al. Safety and Efficacy of the BNT162b2 mRNA Covid-19 Vaccine. New England Journal of Medicine 2020; 383(27): 2603–15.

25. Cheng Y, Ma N, Witt C, et al. Face masks effectively limit the probability of SARS-CoV-2 transmission. Science 2021; 372(6549): 1439–43.

26. Grinshpun SA, Haruta H, Eninger RM, Reponen T, McKay RT, Lee S-A. Performance of an N95 Filtering Facepiece Particulate Respirator and a Surgical Mask During Human Breathing: Two Pathways for Particle Penetration. Journal of Occupational and Environmental Hygiene 2009; 6(10): 593–603.

27. Weber A, Willeke K, Marchioni R, et al. Aerosol penetration and leakage characteristics of masks used in the health care industry. Am J Infect Control 1993; 21(4): 167–73.

28. Cheng Y, Ma N, Witt C, et al. Face masks effectively limit the probability of SARS-CoV-2 transmission. Science 2021: eabg6296.

29. Chang S, Pierson E, Koh PW, et al. Mobility network models of COVID-19 explain inequities and inform reopening. Nature 2021; 589(7840): 82–7.

30. Radonovich LJ, J., Simberkoff MS, Bessesen MT, et al. N95 Respirators vs Medical Masks for Preventing Influenza Among Health Care Personnel: A Randomized Clinical Trial. Jama 2019; 322(9): 824–33.

31. MacIntyre CR, Wang Q, Seale H, et al. A randomized clinical trial of three options for N95 respirators and medical masks in health workers. American journal of respiratory and critical care medicine 2013; 187(9): 960–6.

32. Howard J, Huang A, Li Z, et al. An evidence review of face masks against COVID-19. Proc Natl Acad Sci 2021; 118(4): e2014564118.

33. Prather KA, Marr LC, Schooley RT, McDiarmid MA, Wilson ME, Milton DK. Airborne transmission of SARS-CoV-2. Science 2020; 370(6514): 303–4.

34. Prather KA, Wang CC, Schooley RT. Reducing transmission of SARS-CoV-2. Science 2020; 368(6498): 1422–4.

35. Pöhlker ML, Krüger OO, Förster J-D, et al. Respiratory aerosols and droplets in the transmission of infectious diseases. arXiv [physicsmed-ph] 2021.

36. Chu DK, Akl EA, Duda S, et al. Physical distancing, face masks, and eye protection to prevent person-to-person transmission of SARS-CoV-2 and COVID-19: a systematic review and meta-analysis. The Lancet 2020; 395(10242): 1973–87.

37. Jones NR, Qureshi ZU, Temple RJ, Larwood JPJ, Greenhalgh T, Bourouiba L. Two metres or one: what is the evidence for physical distancing in covid-19? BMJ 2020; 370: m3223.

38. Morawska L, Allen J, Bahnfleth W, et al. A paradigm shift to combat indoor respiratory infection. Science 2021; 372(6543): 689–91.

39. Helleis F, Klimach T, Pöschl U. Vergleich von Fensterlüftungssystemen und anderen Lüftungsbzw. Luftreinigungsansätzen gegen die Aerosolübertragung von COVID-19 und für erhöhte Luftqualität in Klassenräumen. 2021.

40. Peng Z, Bahnfleth W, Buonanno G, et al. Practical Indicators for Risk of Airborne Transmission in Shared Indoor Environments and their application to COVID-19 Outbreaks. medRxiv 2021: 2021.04.21.21255898.

41. Wang CC, Prather KA, Sznitman J, et al. Airborne transmission of respiratory viruses. Science 2021; 373(6558): eabd9149.

42. Lelieveld J, Helleis F, Borrmann S, et al. Model Calculations of Aerosol Transmission and Infection Risk of COVID-19 in Indoor Environments. International Journal of Environmental Research and Public Health 2020; 17(21): 8114.

43. Albert E, Torres I, Bueno F, et al. Field evaluation of a rapid antigen test (Panbio™ COVID-19 Ag Rapid Test Device) for COVID-19 diagnosis in primary healthcare centres. Clinical Microbiology and Infection 2021; 27(3): 472.e7-.e10.

44. Hinds WC, Hinds WC. Aerosol Technology: Properties, Behavior, and Measurement of Airborne Particles: Wiley; 1999.

45. Pöschl U. Atmospheric Aerosols: Composition, Transformation, Climate and Health Effects. Angew Chem Int Ed 2005; 44(46): 7520–40.

46. Seinfeld JH, Pandis SN. Atmospheric Chemistry and Physics, from Air Pollution to Climate Change. New York: John Wiley; 2006.

47. Lelieveld J, Evans JS, Fnais M, Giannadaki D, Pozzer A. The contribution of outdoor air pollution sources to premature mortality on a global scale. Nature 2015; 525: 367.

48. Lelieveld J, Poschl U. Chemists can help to solve the air-pollution health crisis. Nature 2017; 551(7680): 291–3.

49. Zhang R, Li Y, Zhang AL, Wang Y, Molina MJ. Identifying airborne transmission as the dominant route for the spread of COVID-19. Proc Natl Acad Sci 2020; 117(26): 14857–63.

50. Emerging COVID-19 success story: South Korea learned the lessons of MERS. 2021. https://ourworldindata.org/covid-exemplar-south-korea.

51. Emerging COVID-19 success story: Vietnam’s commitment to containment. 2021.

52. COVID-19: Minimisation and protection strategy for Aotearoa New Zealand. 2021.

53. Dehning J, Zierenberg J, Spitzner FP, et al. Inferring change points in the spread of COVID-19 reveals the effectiveness of interventions. Science 2020; 369(6500): eabb9789.

54. Moore S, Hill EM, Tildesley MJ, Dyson L, Keeling MJ. Vaccination and non-pharmaceutical interventions for COVID-19: a mathematical modelling study. The Lancet Infectious Diseases 2021; 21(6): 793–802.

55. Priesemann V, Bodenschatz E, Ciesek S, et al. Nachhaltige Strategien gegen die COVID-19-Pandemie in Deutschland im Winter 2021/2022: Technische Universität Berlin, 2021.

